# Genetic evidence for serum amyloid P component as a drug target for treatment of neurodegenerative disorders

**DOI:** 10.1101/2023.08.15.23293564

**Authors:** A Floriaan Schmidt, Chris Finan, Sandesh Chopade, Stephan Ellmerich, Martin N Rossor, Aroon D Hingorani, Mark B Pepys

**Affiliations:** Institute of Cardiovascular Science, Faculty of Population Health, University College London, London, United Kingdom; UCL British Heart Foundation Research Accelerator, London, United Kingdom; Department of Cardiology, Amsterdam Cardiovascular Sciences, Amsterdam University Medical Centres, University of Amsterdam, Amsterdam, the Netherlands; Department of Cardiology, Division Heart and Lungs, University Medical Center Utrecht, Utrecht University, Utrecht, the Netherlands; Wolfson Drug Discovery Unit, Division of Medicine, University College London, London, United Kingdom; UCL Queen Square Institute of Neurology, Faculty of Brain Sciences, University College London, United Kingdom

**Author notes:** Corresponding author: Amand Floriaan Schmidt,. These authors contributed equally (A Floriaan Schmidt, Chris Finan). **One Sentence Summary:** Genetic analyses of circulating serum amyloid P component (SAP) values suggest that depletion of plasma SAP may decrease the risk of Alzheimer’s disease and Lewy body dementia.

**Keywords:** Alzheimer’s disease, dementia, genome-wide association study, Lewy body dementia, Mendelian randomization, miridesap, neurodegeneration, serum amyloid P component

## Abstract

The direct causes of neurodegeneration underlying Alzheimer’s disease (AD) and many other dementias, are not known. Here we identify serum amyloid P component (SAP), a constitutive plasma protein normally excluded from the brain, as a potential drug target. After meta-analysis of three genome-wide association studies, comprising 44,288 participants, *cis*-Mendelian randomization showed that genes responsible for higher plasma SAP values are significantly associated with AD, Lewy body dementia and plasma tau concentration. These genetic findings are consistent with experimental evidence of SAP neurotoxicity and the strong, independent association of neocortex SAP content with dementia at death. Depletion of SAP from the blood and from the brain, as is provided by the safe, well tolerated, experimental drug, miridesap, may therefore contribute to treatment of neurodegeneration.

## INTRODUCTION

The direct causes and mechanisms of neuronal cell death responsible for the cognitive loss in Alzheimer’s disease (AD) and many other dementias, are not known. Serum amyloid P component (SAP) is an almost invariant, constitutive, normal plasma glycoprotein produced exclusively in the liver. It circulates at a mean (SD) concentration of about 24 (8) mg/l in women and 32 (7) mg/l in men^1^ but it is normally rigorously excluded from the central nervous system (CNS). Cerebrospinal fluid (CSF) concentrations of SAP are one thousand-fold lower than the plasma concentration,^2,3^ presumably reflecting relative impermeability of the blood brain barrier (BBB). There is also evidence for an active transport mechanism exporting SAP from the CSF back into the blood^4^. SAP is named for its universal presence in all human amyloid deposits, which reflects the avid but reversible calcium dependent binding of SAP to all types of amyloid fibrils regardless of their protein composition^5,6^. Thus, although CSF and brain content of SAP are normally extremely low, SAP is nonetheless always present in the intracerebral Aβ amyloid plaques, cerebrovascular Aβ amyloid deposits and the majority of neurofibrillary tau tangles in AD. The binding of SAP stabilises amyloid fibrils^7^ and promotes their formation^8,9^, thereby contributing to amyloid deposition and persistence^10^. Furthermore, accumulation of SAP on intracerebral amyloid plaques, cerebrovascular amyloid deposits and neurofibrillary tangles also increases exposure of cerebral neurones to SAP.

Cerebral Aβ amyloid is a defining feature of AD, is also often present in Lewy body dementia (LBD) and is present in chronic traumatic encephalopathy. It is still not known how amyloid pathology contributes to neurodegeneration but recent reports of cognitive benefit from antibody treatments that reduce the Aβ amyloid burden in AD are encouraging^11,12^. Typical AD neuropathology is often seen in the brains of individuals who were cognitively normal at death, raising the possibility of other pathogenetic factors in dementia. It is therefore interesting that, unrelated to its contribution both to Aβ amyloid formation and persistence, human SAP is itself directly neurotoxic to cerebral neurones *in vitro*^13–16^ and in animal models *in vivo*^17^. Furthermore, neocortical SAP content is significantly associated with dementia at death, independently of neuropathological severity, consistent with a more direct, amyloid-independent, pathogenetic role of SAP in neurodegeneration^18^. Indeed, many of the risk factors for dementia, including cerebral and cerebrovascular amyloid deposition, traumatic brain injury, cerebral haemorrhage and even ‘normal’ ageing, with its associated impairment of the BBB^19^, are characterised by increased exposure of the brain to SAP.

In order to rigorously explore the potential causative role of human SAP in human neurodegenerative diseases, we have now sought genetic epidemiological evidence. SAP is encoded by the gene *APCS* (ENSG00000132703) located on chromosome 1, in close proximity to *CRP* (ENSG00000132693) which encodes C-reactive protein (CRP). These two proteins comprise the pentraxin family, sharing 54% strict residue for residue amino acid sequence homology, even higher genetic sequence homology and having the same secondary, tertiary and quaternary structural organisation. Despite notable phylogenetic conservation of gene and protein sequence and structure among pentraxins, there are marked biological differences between these proteins both within and between species^20^. Thus, human CRP is the classical acute phase protein that is among the most commonly used routine clinical chemistry analytes, whilst human SAP is a constitutive plasma protein, the assay of which has hitherto had no practical clinical significance. Human SAP is not an acute phase reactant although in chronic inflammatory conditions, in which there is sustained increased production of CRP, SAP values tend to be slightly higher, albeit within the reference range^21^. A few small studies in the elderly and subjects with impaired cognition have reported plasma and CSF SAP concentrations above the reference range of the healthy middle aged population^2,22,23,24^. Children under 10 years have circulating SAP concentrations below the adult range but reduced adult SAP values are seen only with severe hepatocellular impairment^21^. Unsurprisingly therefore, in contrast to CRP concentration, there have only been limited genome-wide association studies (GWAS) of plasma SAP concentration^24–26^. Recently, however, the SomaLogic aptamer-based proteomic platform has enabled large scale measurement of circulating SAP abundance, allowing for a growing number of GWAS identifying potential genetic instruments for plasma SAP concentration.

*Cis*-Mendelian randomization (MR) leverages genetic instruments associated with protein concentration to demonstrate the possible causal effects of a potential drug target and thus to anticipate safety and efficacy outcomes of specific therapeutic interventions. The random allocation of genes during gametogenesis crucially protects genetic associations against bias due to confounding and reverse causality^27,28^. Furthermore, through a two-sample design, MR can source aggregated data, that is point estimates and standard errors, from large scale studies, each designed to maximize the available sample size. This extensively validated MR approach can provide a precise and powerful overview of the likely causal consequences of target perturbation covering a large number of clinically relevant diseases and traits^29–32^.

We confirmed that the SomaLogic SAP values reliably reflect the actual plasma concentration of the protein measured by rigorously calibrated SAP immunoassay. We then conducted a meta-analysis of three GWAS of circulating SAP values, combining information from 44,288 participants, followed by a drug target MR utilizing *APCS cis*-variants that were strongly associated with plasma SAP values. We primarily focused on the possible causal effect of SAP in AD and LBD. Given the close proximity of *APCS* to *CRP*, and the major involvement of CRP responses with almost all inflammatory, infective, traumatic and other tissue damaging processes^33^, we additionally utilized MR to rule out possible effects of plasma CRP concentration acting on the SAP signal through linkage disequilibrium (LD) between variants in *APCS* and variants in *CRP*.

## RESULTS

### Genome-wide meta-analysis of plasma SAP values

Genetic variant-specific estimates of the association with SAP values, measured by SomaLogic SomaScan assay version 4.1, were available from three independent studies: Interval, comprising 3,301 particpants^24^, AGES, with 5,368^26^, and DECODE with 35,559^25^. The combined data identified ten independent lead variants associating with SAP, including four *cis*-variants near *APCS* (rs140308485, rs13374652, rs1341664, rs78228389), as well as *trans*-variants on chromosomes 1, 2, 8 and 13 (Fig. 1-2, Table S1). Comparison of the genetic associations with plasma SAP and CRP values in the region around rs1341664, the *APCS cis*-variant with the strongest SAP association, indicated that these closely adjacent signals were independent (Fig. 2); the Pearson correlation coefficient comparing the -log_10_(p-value) for each trait was -0.06, p-value < 0.001. This was further confirmed by noting that the CRP association of the four SAP *cis*-variants did not reach genome-wide significance (Table S2), with the -log_10_(p-value) for CRP ranging between 0.04 and 6.26. Instead, the CRP signals were concordant with the SAP *trans* signals (Fig. S2; Table S2).

**Fig. 1.**
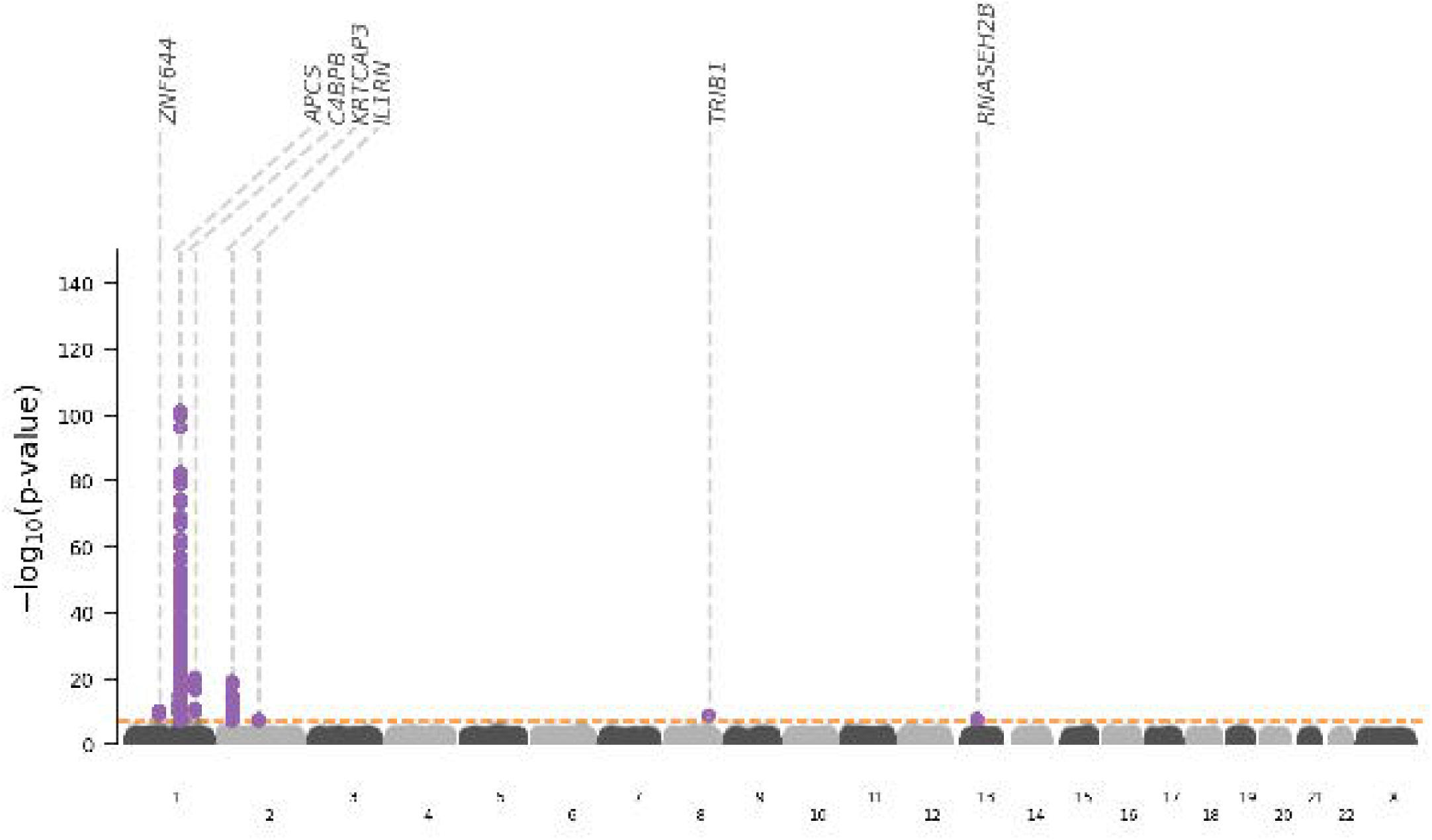
Manhattan plot of SAP genome-wide association study. The -log_10_(p-value) of genetic variants is shown on the y-axis and GRCh37 base pair position within chromosomes on the x-axis. The horizontal dashed line is at p-value 5.8×10^-8^. The lead variants are labelled with the putative causal genes assigned by V2G.

**Fig. 2.**
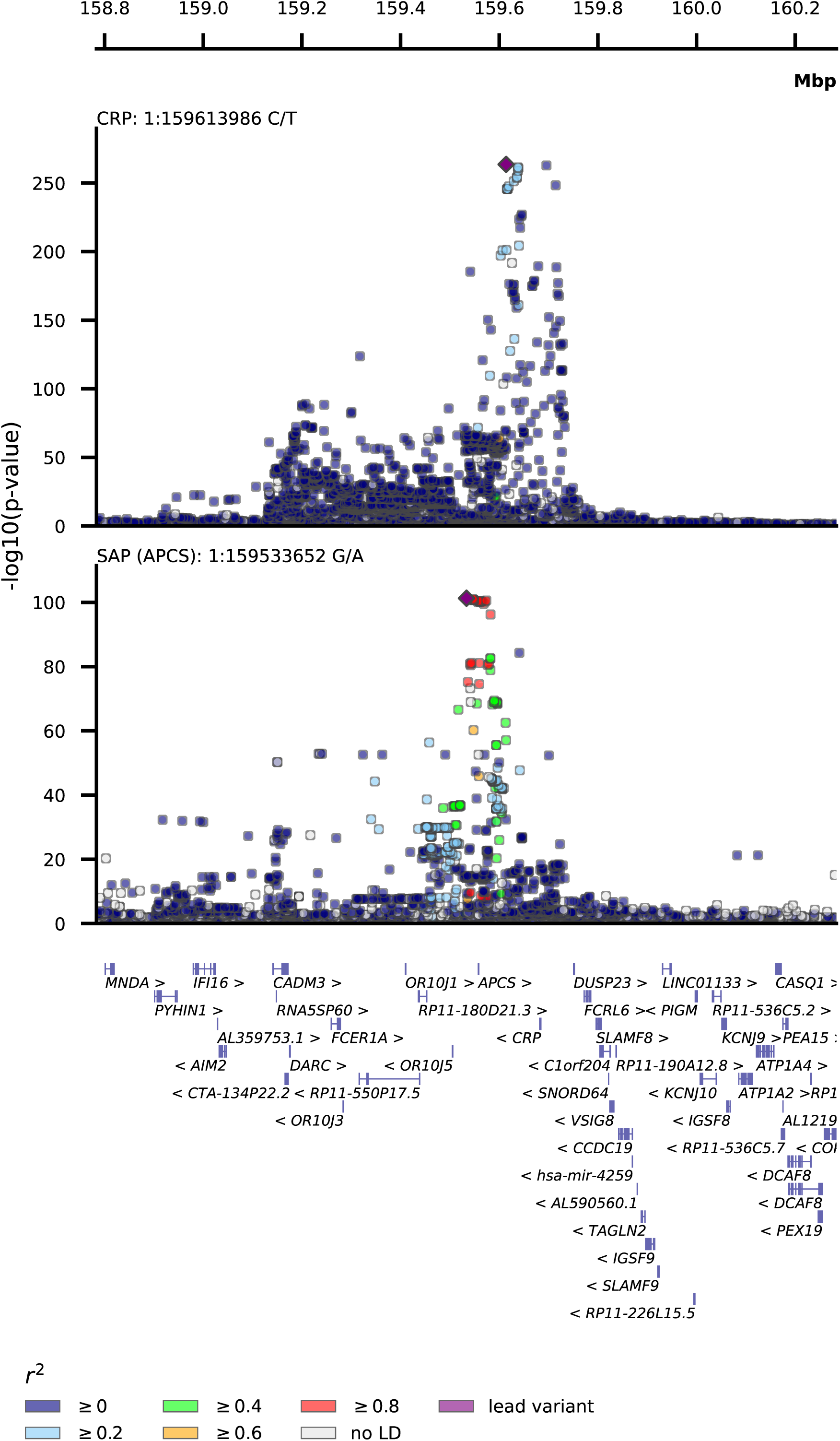
Stacked locus-view comparing the overlap between genetic variants for plasma SAP values and plasma CRP concentration. The -log_10_(p-value) of the genetic association with SAP and CRP plotted (y-axis) against the genomic location (x-axis). The lead variant for each trait is indicated by a purple diamond. Linkage disequilibrium with the lead variant is indicated by coloured dots, with the r-squared estimated from the UKB and 1000 genomes EUR reference. Gene locations were queried from Ensembl v109 (GRCh37).

**Fig. 3.**
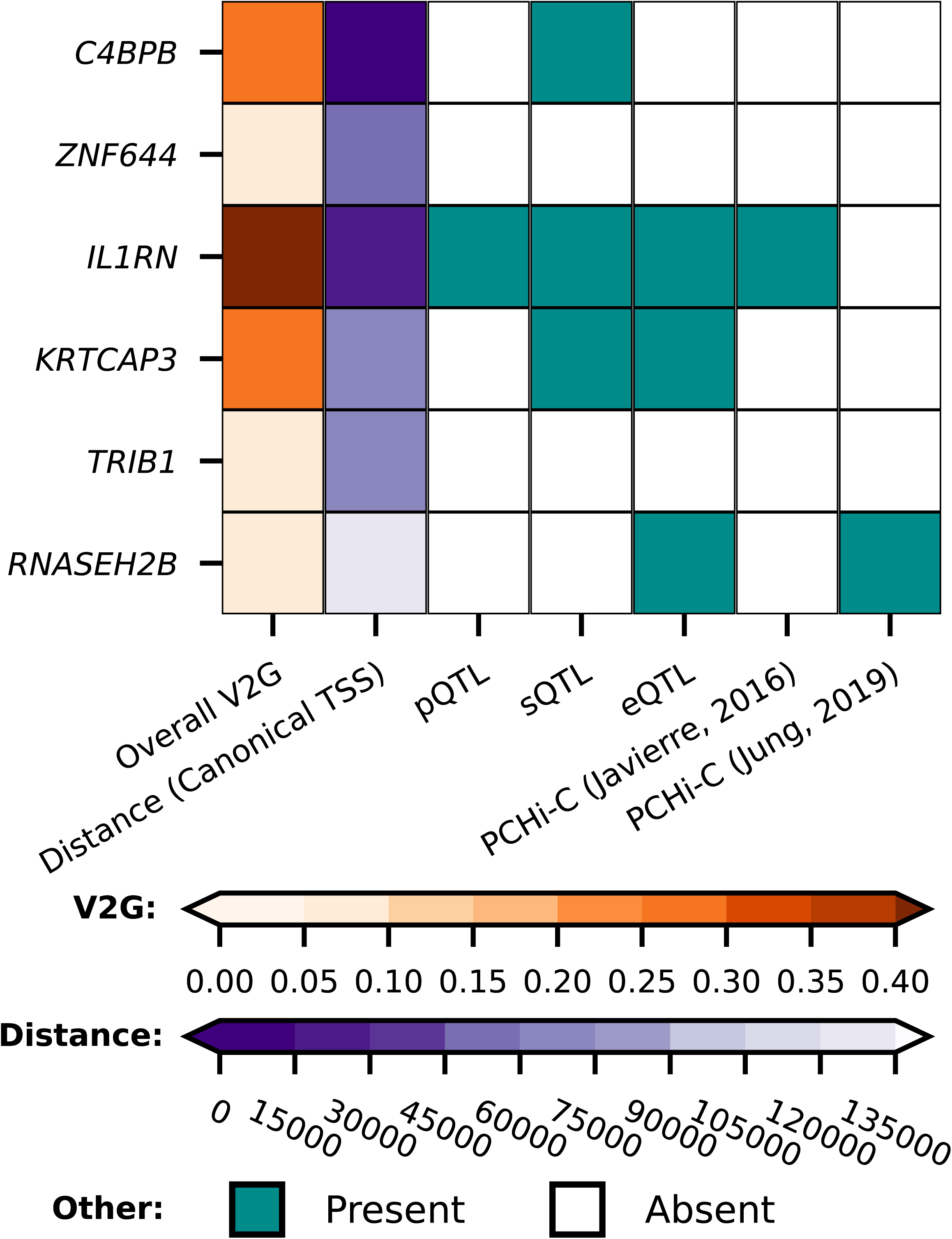
Mapping of the *trans* signals for variants associated with plasma SAP value to putative causal genes. First column, the overall V2G score; second column, distance in base pairs from the lead variant to the canonical gene transcription start site (TSS). The other columns show the presence or absence of x-axis criteria, specifically, whether there were quantitative trait loci (QTL) linking the gene to proteomics, transcriptomics, or splice site QTL, and PCHi-C (Promoter Capture Hi-C) chromatin interaction experiments linking the genetic variant to the indicated gene, from Jung *et al.*^65^ and Javierre *et al.*^66^. Tables S3-S8 show the whole V2G output.

GWAS Catalog look-ups for the effects of genetic variants in *APCS* identified a previously reported association with white-matter microstructure^34^ (Fig. S3; Table S9). Using Open Target’s V2G algorithm the six SAP *trans*-variants were mapped to the putative causal genes: *C4BPB, ZNF644, IL1RN, KRTCAP3, TRIB1, RNASEH2B* (Fig. 1; 3, Tables S3-S8). Look-ups for the variants within and around the putative *trans*-genes for SAP, provided links with a diverse range of pathophysiology without known connections to SAP biology (Fig. S3, Table S9).

### Cis-MR results for plasma SAP values and dementia outcomes

*Cis*-MR analysis detected significant associations of higher plasma SAP values with increased risk of dementia outcomes: AD (35,274 cases, odds ratio (OR) 1.07, 95%CI 1.02; 1.11, p=1.8×10^-3^), and LBD (2,981 cases, OR 1.37, 95%CI 1.19; 1.59, p=1.5×10^-5^) (Fig. 4, Table S10). A similar analysis for plasma CRP values did not identify links with these outcomes (Table S10).

**Fig. 4.**
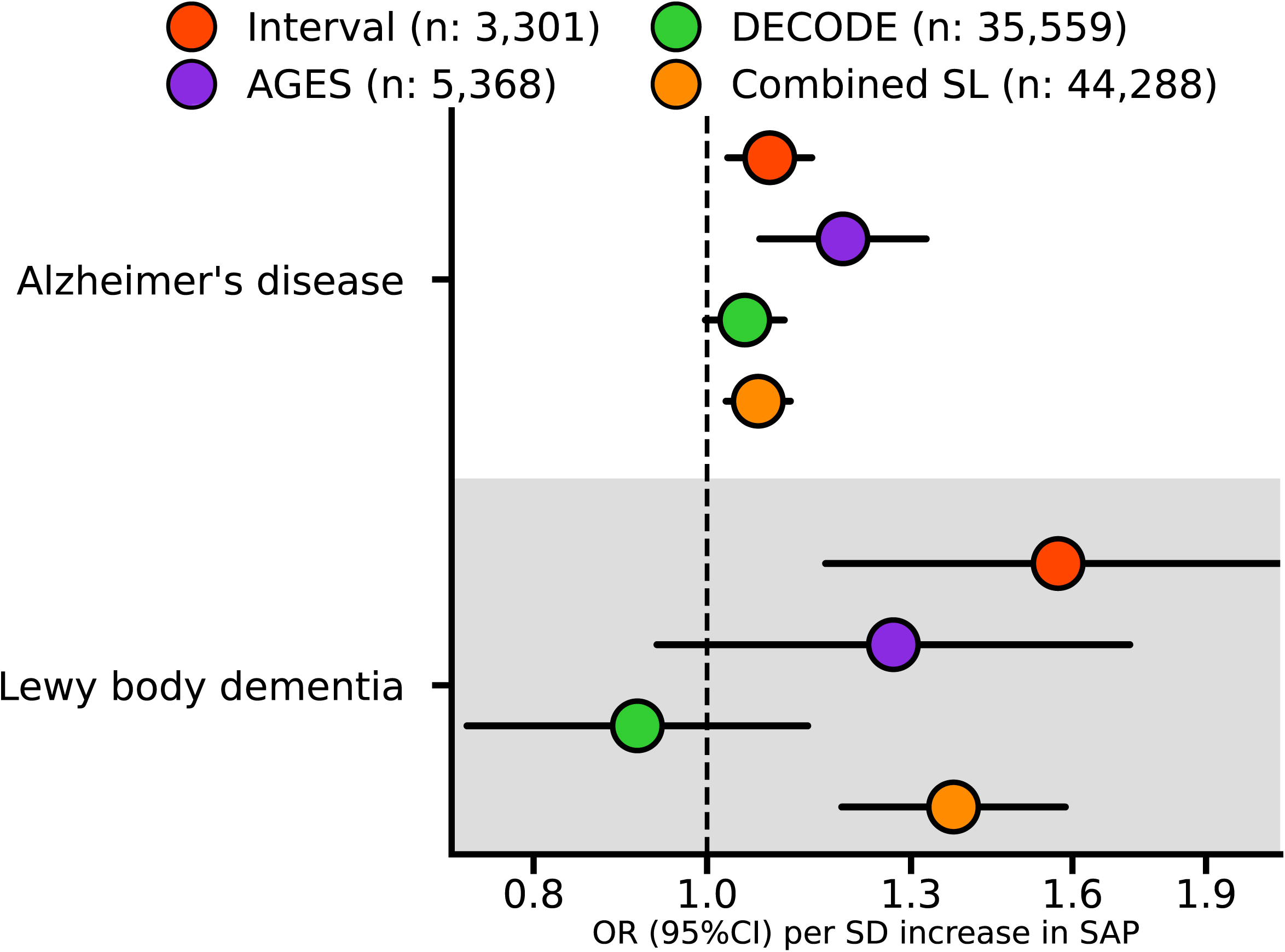
Estimates of the *cis* Mendelian randomization effect of plasma SAP values on Alzheimer’s disease and Lewy body dementia. The univariable MR effects estimated from the GWAS meta-analysis of SomaLogic (SL) plasma SAP values from the Interval, AGES and DECODE studies are shown individually, together with the combined result of all three studies. We used the AD GWAS from Kunkle *et al.*^52^ which consisted of 35,274 cases and 59,163 controls, the LBD GWAS from Chia *et al.*^53^ consisted of 2,981 cases and 2,173 controls. OR, odds ratio; 95%CI, 95% confidence interval. The Figure illustrates the data in Table S10.

### Cis-MR results for plasma SAP values and other outcomes

*Cis-*MR analysis suggested that higher plasma SAP value was associated with increased coronary heart disease (CHD) risk (OR 1.03, 95%CI 1.01; 1.05), greater total brain volume (0.06 SD, 95%CI 0.02; 0.10), lower systolic blood pressure (SBP) (-0.16 mm Hg, 95%CI -0.26; -0.07) and lower diastolic blood pressure (DBP) (Fig. 5, Table S11). In contrast, the MR analysis of plasma CRP values showed a distinct effects profile in which higher CRP concentrations were associated with serum concentrations of hepatocellular enzymes, with osteoarthritis, and with total plasma tau concentration (Fig. 5, Table S12).

**Fig. 5.**
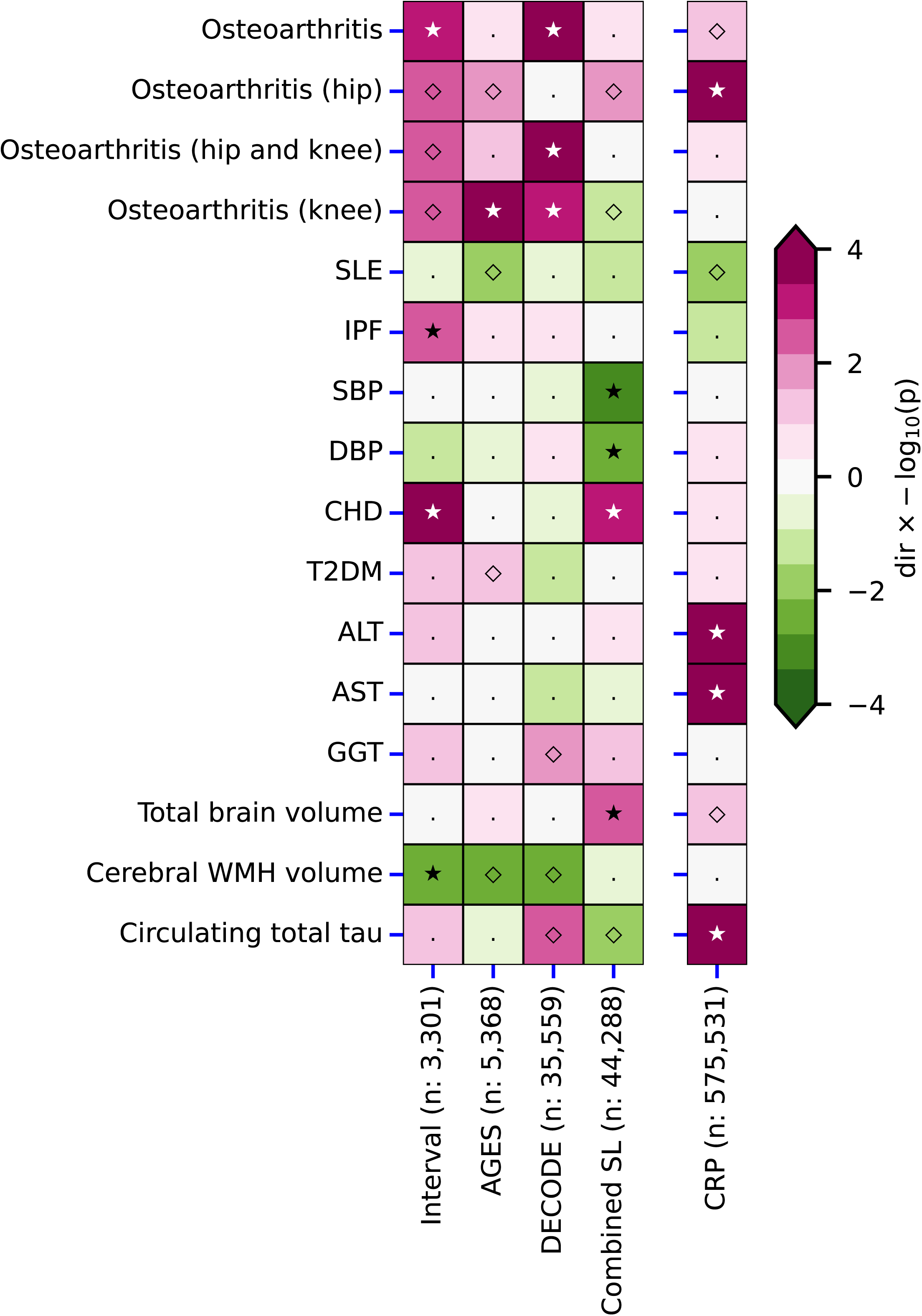
Comparison of the *cis* Mendelian randomization effect estimates of plasma SAP values and CRP concentration. Effect direction, magenta for positive and green for negative, is shown by the -log10(p-value) to 4 significant figures. Open diamonds, p<0,05; closed stars, p<2.38 x 10^-5^. Left block, SAP; right column, CRP. Abbreviations: ALT, alanine transaminase; AST, aspartate transaminase; CHD, coronary heart disease; DBP, diastolic blood pressure; GGT, gamma-glutamyl transferase; IPF, idiopathic pulmonary fibrosis; SBP, systolic blood pressure; SLE, systemic lupus erythematosus; T2DM, type 2 diabetes; WMH, white matter hyperintensities. The Figure illustrates the data in Tables S11-S12.

### Multivariable MR results for outcomes linked to both SAP and CRP

*APCS* and *CRP* are closely co-located on chromosome 1, potentially challenging the recognition of, and discrimination between, more subtle apparent independent effects. We therefore identified outcomes significant at the more liberal value of p<0.05 for both plasma SAP and CRP values, specifically, total brain volume, plasma total tau concentration and osteoarthritis. Application of multivariable MR (MVMR) to identify the mutually independent effects of both proteins, and accounting for potential influence of LD, then showed that higher values of each increased circulating total tau concentration: (0.06 log_2_(ng/L), 95%CI 0.03; 0.08) for higher plasma SAP value, and (0.20 log_2_(ng/L), 95%CI 0.14; 0.25) for higher plasma concentration of CRP. The MVMR analysis did not confirm significant associations for osteoarthritis, and total brain volume (Table S13).

## DISCUSSION

We report here a large-scale meta-analysis of GWAS of SomaLogic values for plasma SAP comprising 44,288 participants. We confirmed that the SomaLogic intensity scores for plasma SAP agree with immunoassay of actual SAP concentrations. The GWAS results then enabled MR analysis sourcing *cis*-acting variants within and around *APCS*, the gene encoding SAP, to explore potential involvement of SAP in pathogenesis of dementia. We found that higher plasma SAP values increased the risk of AD (OR 1.07, 95%CI 1.02; 1.11, p=1.8×10^-3^) and LBD (OR 1.37, 95%CI 1.19; 1.59, p=1.5×10^-5^), implying that pharmaceutical depletion of SAP might reduce the risk of both diseases. Furthermore, using multivariable MR to account for possible horizontal pleiotropy by plasma CRP concentration, we also detected a significant association of higher plasma SAP values with higher total plasma tau concentration (0.06 log_2_(ng/L), 95%CI 0.03; 0.08), which is itself also associated with AD dementia^35^.

The presence of SAP in all cerebral Aβ plaques and on most neurofibrillary tangles in AD, long known from immunohistochemical studies, has recently been shown to strongly discriminate between AD brains and cognitively normal brains^36^. However, the present finding of significant associations between genetically determined higher plasma SAP values and increased risk of AD and LBD, is more specifically consistent with the association between neocortex SAP content and cognitive status at death that was recently observed in the Cognitive Function and Ageing Study^18^. In this unselected, population-representative, elderly brain donor population, the OR for dementia at death, between the top tertile and the lowest tertile of neocortical SAP content, was 5.24 (95%CI 1.79; 15.29). Furthermore the association of dementia with SAP content was independent of Braak stage, Thal phase and all other classical neuropathological hallmarks of dementia^18^. It was thus specific for abundance of SAP itself in the neocortex rather than SAP content just being a surrogate for the Aβ amyloid and neurofibrillary tau tangle pathology which is always present in AD and also frequently found in LBD. This SAP-dementia association, which is consistent with a possible pathogenetic role of SAP in neurodegeneration, is now directly supported by the present MR results.

In contrast to a previously reported MR study, which did not detect an association between SAP values and AD^37^, our analysis was improved in multiple ways. First and foremost, instead of simply taking a single SAP GWAS, we meta-analysed combined data from three independent studies to produce the largest GWAS of SAP values to date. This resulted in an AD MR analysis using 53 SAP variants instead of the 14 used by Yueng *et al*^37^. The larger number of variants allowed us to consider outlier and leverage statistics, identifying and removing variants with possible horizontal pleiotropic effects, further ensuring the robustness of the present findings. Furthermore, we performed confirmatory analyses to refute possible bias due to the location of the *CRP* gene closely adjacent to *APCS*, and we found no meaningful overlap between the effect profiles of plasma SAP and CRP values. Similar to the analysis by Yueng *et al*^37^, we conducted a two-sample MR analysis, where the exposure GWAS did not, or only partially, overlapped with the outcome GWAS. Any potential weak instrument bias will thus, on average, act towards a null effect, hence our results are conservative. This, however, also implies that non-significant findings should not be over-interpreted as providing proof of absence^38^. Finally, we note that, under the null hypothesis and accounting for the number of evaluated outcomes, the probability of finding an effect of plasma SAP value on AD as well as on LBD is equal to the square of the alpha: (2.78×10^-3^)^2^ =7.73×10^-6^. The concordant findings thus strongly imply that SAP contributes to pathogenesis of dementia.

Increased duration and/or intensity of brain exposure to SAP may be pathogenic through its direct cytotoxicity for some cerebral neurones, as has been demonstrated experimentally *in vitro* and *in vivo*^13–16^, and potentially also by promoting the formation and the persistence of Aβ amyloid fibrils and neurofibrillary tau tangles. However, SAP is produced only by the liver; it is not in the brain transcriptome^39^. Normally the brain is strongly protected against exposure to SAP by the BBB and by active transport back to the plasma of any SAP that leaks through^4^, so that the CSF SAP concentration is about one thousandth of that in the plasma^2,3^. It is therefore striking that even modestly higher plasma SAP concentrations are associated with dementia risk. There have been very few studies of SAP concentration in paired samples of plasma or serum and CSF but in addition to the circulating SAP concentration, BBB integrity and the efficiency of the SAP active export mechanism must affect brain exposure to SAP. Nevertheless, across the large populations studied here, there was a significant effect of higher plasma SAP concentration on clinical dementia outcomes.

In addition to the four *cis*-loci, our GWAS results identified variants mapping to genes outside *APCS*, including *C4BPB, ZNF644, IL1RN, KRTCAP3, TRIB1* and *RNASEH2B*. SAP binds specifically to C4-binding protein, encoded by *C4BPB*, under particular experimental conditions *in vitro*^40^, though no functional effect of the interaction has been reported. *IL1RN*, which encodes the IL-1 receptor antagonist (IL-1RA) might have a functional effect on SAP via the acute phase response, which is mediated by IL-1 both directly and via other pro-inflammatory cytokines. Even though human SAP is not an acute phase reactant, its concentration does tend to rise modestly within the reference range in chronic inflammatory diseases with a sustained acute phase response^21^. *TRIB1* has diverse, wide ranging effects across many different physiological systems.

The putatively mapped SAP *trans-*genes have previously been variously linked to a broad range of different metabolic, cardiac and haematological traits and to increased plasma concentrations of liver enzymes. Inclusion of these traits in our *cis*-MR analysis identified an association of higher SAP values with increased CHD and decreased SBP and DBP, but, since there is no known functional connection between these cardiovascular features and SAP, the protein itself is unlikely to be directly involved. Potential pathogenetic connections have been suggested between SAP and two different, unrelated diseases, osteoarthritis^41^ and systemic lupus erythematosus^42^. Plasma SAP concentrations were not strongly associated with lupus but there were apparent associations between increased SAP values and some osteoarthritis outcomes (Fig. 5), perhaps reflecting our conservative analyses. We also found a significant positive association between SAP and idiopathic pulmonary fibrosis, suggesting that SAP may also have a pathogenetic role in this condition (Fig. 5). In contrast, osteoarthritis^43^ and lupus^44^, respectively, have well established positive and negative clinical links with CRP, and, interestingly, both were associated with circulating CRP concentrations in the corresponding directions in the current analysis (Fig. 5).

Potential limitations to our study comprise, firstly, our use of SomaLogic values for plasma SAP, which are only relative intensities not actual SAP concentrations. SomaLogic assays alone therefore cannot enable precise determination of the effect magnitude relevant for potential pharmaceutical intervention, even though we rigorously demonstrated that the SomaLogic values are in acceptably close agreement with the actual SAP concentrations measured by precise, rigorously standardised electroimmunoassay.

Secondly, previous GWAS studies of AD and other types of dementia have not reported *APCS* as a potential gene for disease onset. However, GWAS is deliberately designed to limit detection of false positive results and may accordingly leave additional signals undiscovered^27^. It is therefore important to emphasize that drug target MR does not require the GWAS data used for the outcome trait to reach GWAS significance^28^.

Thirdly, while the present results imply that SAP depletion might reduce dementia risk, they do not indicate optimal timing for the intervention. Neuropathological changes are well known to long precede clinically detectable cognitive loss in AD and other dementias, so SAP depletion might be most effectively introduced prophylactically. Nevertheless, in view of the present evidence for a causal relationship between increased circulating SAP and risk of dementia, prompt SAP depletion may protect residual cognition at any stage.

Fortunately, the experimental drug, miridesap, (CPHPC; hexanoyl bis-D-proline; (R)-1-[6-[(R)-2-carboxy-pyrrolidin-1-yl]-6-oxo-hexanoyl]pyrrolidine-2-carboxylic acid))^45^, safely provides extremely effective SAP depletion. It reduces plasma SAP concentration by more than 95% for as long as the drug is administered^46^ and thereby removes all detectable SAP from the CSF in patients with AD^3^ and from the brain in human SAP transgenic AD model mice^47^. DESPIAD, a small, academic, phase 2b clinical trial of SAP depletion by miridesap in established AD, is now in progress (EudraCT number 2016-003284-19) and will report in 2025. Meanwhile, our present genetic analysis indicates that depletion of plasma SAP is expected to decrease the risk of AD and LBD.

## MATERIALS AND METHODS

### Validation of SomaLogic SAP assay

The read out from the SomaLogic aptamer-based mass spectrometric method is relative reagent intensity, a proxy for SAP concentration rather than actual mass per volume. We therefore used the robust electroimmunoassay method, rigorously standardised with isolated, pure human SAP^1^, to measure the actual concentration of SAP in 100 human plasma samples from a random sub-cohort of the EPIC-Norfolk study (https://www.epic-norfolk.org.uk/) in which SAP had been quantified by the SomaLogic method. The electroimmunoassay confirmed that the SomaLogic results accurately reflected plasma SAP concentrations: for the two sets of results, the Pearson correlation coefficient was 0.86 (p< 0.001) and Spearman correlation was 0.84 (p<0.001) (Fig. S1).

### GWAS of SAP plasma values

Three independent studies with plasma SAP values determined by the SomaLogic method: Interval (n: 3,301)^24^, AGES (n: 5,368)^26^, and DECODE (n: 35,559)^25^, were used to provide aggregate genetic data. To account for potential heterogeneity in genetic associations, due to difference in participants and/or environment, we performed a DerSimonian-Laird random effects meta-analysis using METAL^48^.

Independent lead genetic variants were identified by filtering associations on a genome-wide significant p-value of 5.8×10^-8^ and clumping to an R-squared of 0.01 based on LD reference data from a random 5,000 participant subset of the UK Biobank (UKB). The nearest protein coding genes were identified by querying the GRCh37 assembly via Ensembl REST API^49^. Lead variants within 2 megabase pairs (MB) of *APCS*, the gene encoding SAP, were assigned to this gene; *trans-*variants (outside ±2 MB of *APCS*) were mapped to putative causal genes using the V2G algorithm offered by Open Targets^50^. The V2G algorithm ranks putative causal genes based on integrated information on molecular traits, such as, information on splice-sites, mRNA expression, chromatin interaction, functional predictions and distance from the canonical transcription start site. Potential pleiotropic associations of these putative causal genes were explored by querying the author-assigned gene in GWAS Catalog, which comprises the largest source of gene to phenotype information^51^.

### Drug target Mendelian randomization

Drug target MR was employed to ascertain the possible causal effects that a unit increase in standard deviation (SD) of mean plasma SAP concentration had on clinically relevant traits, with a primary focus on AD^52^ and LBD^53^.

To limit the potential for bias due to pre-translational horizontal pleiotropy, variants were extracted from within and around *APCS*, applying a ±1 MB pairs flank^54^. Variants were filtered to a minor allele frequency of 0.01 or larger, and clumped to an R-squared of 0.40. Residual LD was modelled using generalized least square (GLS) solutions^55^ and a 5,000 random sample of UKB participants. To reduce the risk of weak-instrument bias^56^, we selected genetic variants with an F-statistic of 15 or higher. Furthermore, due to the absence of sample overlap between the SAP GWAS dataset and the GWAS used for many of the outcome traits, any potential weak-instrument bias would act towards a null effect, reducing power rather than increasing type 1 errors^56,57^.

Estimates of the potential causal effect of higher plasma SAP value were obtained using the GLS implementation of the inverse-variance weighted (IVW) estimator and the MR-Egger estimator, the latter being unbiased in the presence of horizontal pleiotropy at the cost of lower precision^58^. To minimize the potential influence of horizontal pleiotropy, variants beyond 3 times the mean leverage or an outlier Chi-square statistic larger than 10.83, were pruned^59^. Finally, a model selection framework was applied to select the most appropriate estimator, IVW or MR-Egger^59,60^. This model selection framework^61^ utilizes the difference in heterogeneity between the IVW Q-statistic and the Egger Q-statistic to decide which method provides the best model to describe the available data and hence optimizes the bias-variance trade-off.

Given the close proximity of *CRP* to *APCS* we additionally conducted an MR analysis of CRP concentration, taking advantage of availability of the largest CRP GWAS conducted to date^62^. The MR effect estimates of SAP and CRP were compared to identify outcomes which seemingly were affected by both proteins using a p-value of 0.05. For the subset of outcomes which were affected by both SAP and CRP, we additionally conducted MVMR to analytically control any influence of CRP on the SAP signal and vice versa. MVMR is similar to standard multiple regression, where multiple variables, in our case two, are included in the same model, resulting in estimates that are mutually independent of one another^63^. Importantly, MVMR allowed us to account for any horizontal pleiotropy that might act through CRP concentration^63^. In addition, to correct for any potential remaining horizontal pleiotropy acting through non-CRP pathways, we applied the same model selection framework to decide between MVMR with and without Egger correction. Where relevant, we differentiate between MVMR and regular MR results by referring to the latter as univariable MR.

### Effect estimates and multiple testing

Unless otherwise specified, all point estimates, that is OR or mean differences, refer to a unit change of the independent variable, typically one standard deviation in plasma protein value for MR results or an increase in number of risk alleles for GWAS results, respectively. Results are provided with 95% confidence intervals (CI) and p-values. Significance in the GWAS analysis was evaluated using the standard multiplicity corrected alpha, that is the false positive rate, of 5.8×10^-8^, accounting for the estimated number of independent genetic variants in the genome^64^. The MR results were tested against a Bonferroni corrected alpha of 2.78×10^-3^, accounting for the 18 evaluated traits (Data availability section).

## Acknowledgments

A preprint version of this manuscript has been deposited at medRxiv. We thank Professor Nick Wareham and the EPIC-Norfolk team for generous provision of the plasma samples we used to validate the SomaLogic method for quantification of SAP concentration.

## Author contributions

AFS, CF, MBP conceived and designed the study. AFS, CF, SC performed the analyses. SE ran the immunoassays for SAP. AFS and MBP drafted the manuscript with critical input from CF, SC, SE, ADH and MNR.

## Ethics declaration

The study exclusively uses information from aggregate data resources. As such this study is exempt from approval by an Ethics Committee or Institutional Review Board.

## Data and materials availability

All of the source data for this study are publicly available: SAP plasma value from DECODE (https://download.decode.is/form/folder/proteomics), AGES (https://doi.org/10.5281/zenodo.5711426), and Interval (https://www.ebi.ac.uk/gwas/studies/GCST90242796). Data on CRP concentration were obtained from: https://www.ebi.ac.uk/gwas/studies/GCST90029070. GWAS data were accessed for the following traits: Alzheimer’s disease (https://www.niagads.org/datasets/ng00075), Lewy body dementia (https://www.ebi.ac.uk/gwas/publications/33589841), osteoarthritis (https://www.ebi.ac.uk/gwas/publications/30664745), systemic lupus erythematosus (https://www.ebi.ac.uk/gwas/publications/26502338), idiopathic pulmonary fibrosis, (https://www.ebi.ac.uk/gwas/publications/33197388), systolic/diastolic blood pressure (https://www.ebi.ac.uk/gwas/publications/30224653), coronary heart disease (https://www.ebi.ac.uk/gwas/publications/36474045), type 2 diabetes (http://diagram-consortium.org/downloads.html, from Mahajan *et al.*), liver enzymes (https://www.ebi.ac.uk/gwas/publications/33972514 and https://www.ebi.ac.uk/gwas/publications/33547301), brain volume (https://ctg.cncr.nl/software/summary_statistics, from Jansen *et al*.), cerebral white matter hyperintensities (https://www.ebi.ac.uk/gwas/publications/26674333), circulating total tau values (https://www.ebi.ac.uk/gwas/publications/35396452). All data needed to evaluate the conclusions in the paper are present in the paper and the Supplementary Materials.

## List of Supplementary Materials

### Figures

S1 Correlation between SomaLogic values and immunoassay for actual plasma SAP concentration

S2 Locus view plots

S3 GWAS Catalog look-upsTables

S1 Lead variants for serum amyloid P component (SAP) plasma value

S2 SAP lead variant associations with plasma C-reactive protein (CRP) concentration

S3 Open target v2g results mapping *trans*-variant rs2808467 to a putative causal gene

S4 Open target v2g results mapping *trans*-variant rs165316 to a putative causal gene

S5 Open target v2g results mapping *trans*-variant rs10188292 to a putative causal gene

S6 Open target v2g results mapping *trans*-variant rs4665972 to a putative causal gene

S7 Open target v2g results mapping *trans*-variant rs112875651 to a putative causal gene

S8 Open target v2g results mapping *trans*-variant rs9591359 to a putative causal gene

S9 GWAS Catalog look-ups for putative causal genes for plasma SAP value

S10 The *cis*-Mendelian randomization results for the effects of one standard deviation higher plasma SAP value or plasma CRP concentration on dementia outcomes

S11 The *cis*-Mendelian randomization results for the effects of one standard deviation higher plasma SAP value on secondary outcomes

S12 The *cis*-Mendelian randomization results for the effects of one standard deviation higher plasma CRP concentration on secondary outcomes

S13 The *cis*-multivariable Mendelian randomization results for the effects of one standard deviation higher plasma SAP value or plasma CRP concentration

