## Supplementary for "Genetic evidence for serum amyloid P component as a drug target for treatment of neurodegenerative disorders"

Supplementary information

Schmidt *et al*.

S3 Open target v2g results mapping *trans*-variant rs2808467 to a putative causal gene . 13
S4 Open target v2g results mapping *trans*-variant rs165316 to a putative causal gene . . 14
S5 Open target v2g results mapping *trans*-variant rs10188292 to a putative causal gene . 15
S6 Open target v2g results mapping *trans*-variant rs4665972 to a putative causal gene . 16
S7 Open target v2g results mapping *trans*-variant rs112875651 to a putative causal gene 17
S8 Open target v2g results mapping *trans*-variant rs9591359 to a putative causal gene . 18
S9 GWAS Catalog look-ups for putative causal genes for plasma SAP value. . . . 19
S10 The *cis*-Mendelian randomization results for the effects of one standard deviation higher plasma SAP value or plasma CRP concentration on dementia outcomes. . . . 20
S11 The *cis*-Mendelian randomization results for the effects of one standard deviation higher plasma SAP value on secondary outcomes . . . 21
S12 The *cis*-Mendelian randomization results for the effects of one standard deviation higher plasma CRP concentration on secondary outcomes . . . 23
S13 The *cis*-multivariable Mendelian randomization results for the effects of one standard deviation higher plasma SAP value or plasma CRP concentration. . . . 24

### Figures

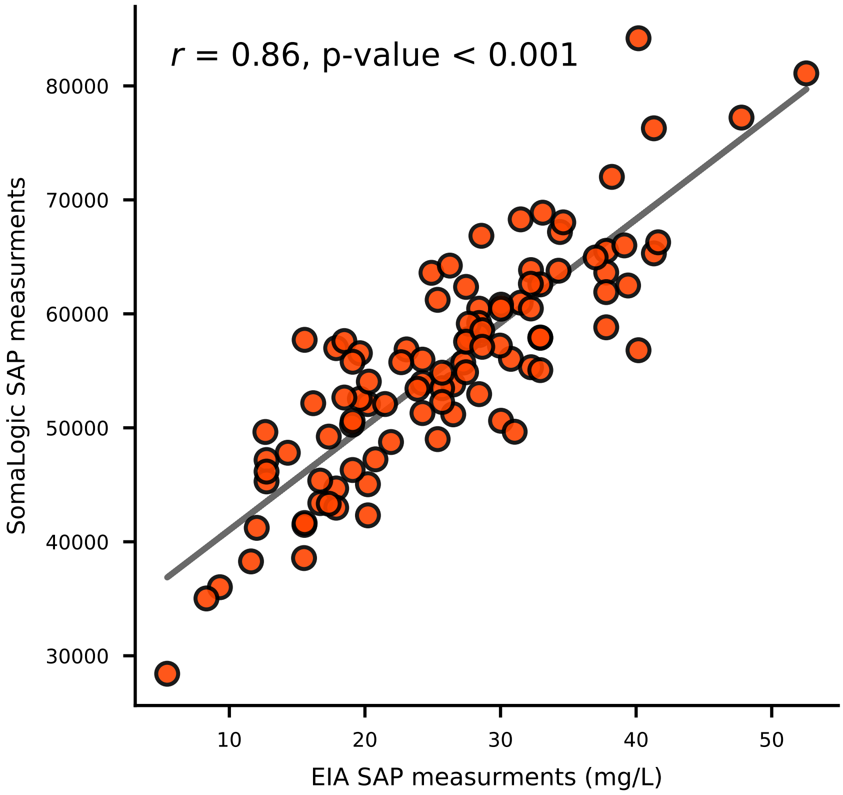

**Figure S1:** The Pearson correlation estimated in 100 plasma samples from the EPIC-Norfolk study, between SomaLogic relative intensity scores for plasma SAP value and electroimmunoassay (EIA) of actual SAP concentration, standardised and calibrated with authentic, pure, comprehensively characterized, native, human SAP.

#### Figure S2:

Locus view plots showing the plasma SAP and CRP signals of the ten independent lead variants identified in the present SAP GWAS meta-analysis of three independent pQTL GWAS based on SomaLogic assays. The variant-specific $-\log_{10}(\text{p-value})$ is plotted (y-axis) against the genomic location (x-axis). The lead variant is indicated by a purple diamond, with linkage disequilibrium (LD) relative to the lead variant indicated by the colours of the dots. The R-squared, representing LD, was from a random sample of 5,000 UK Biobank participants supplemented by the 592 participants of the 1,000 Genomes EUR reference panel.

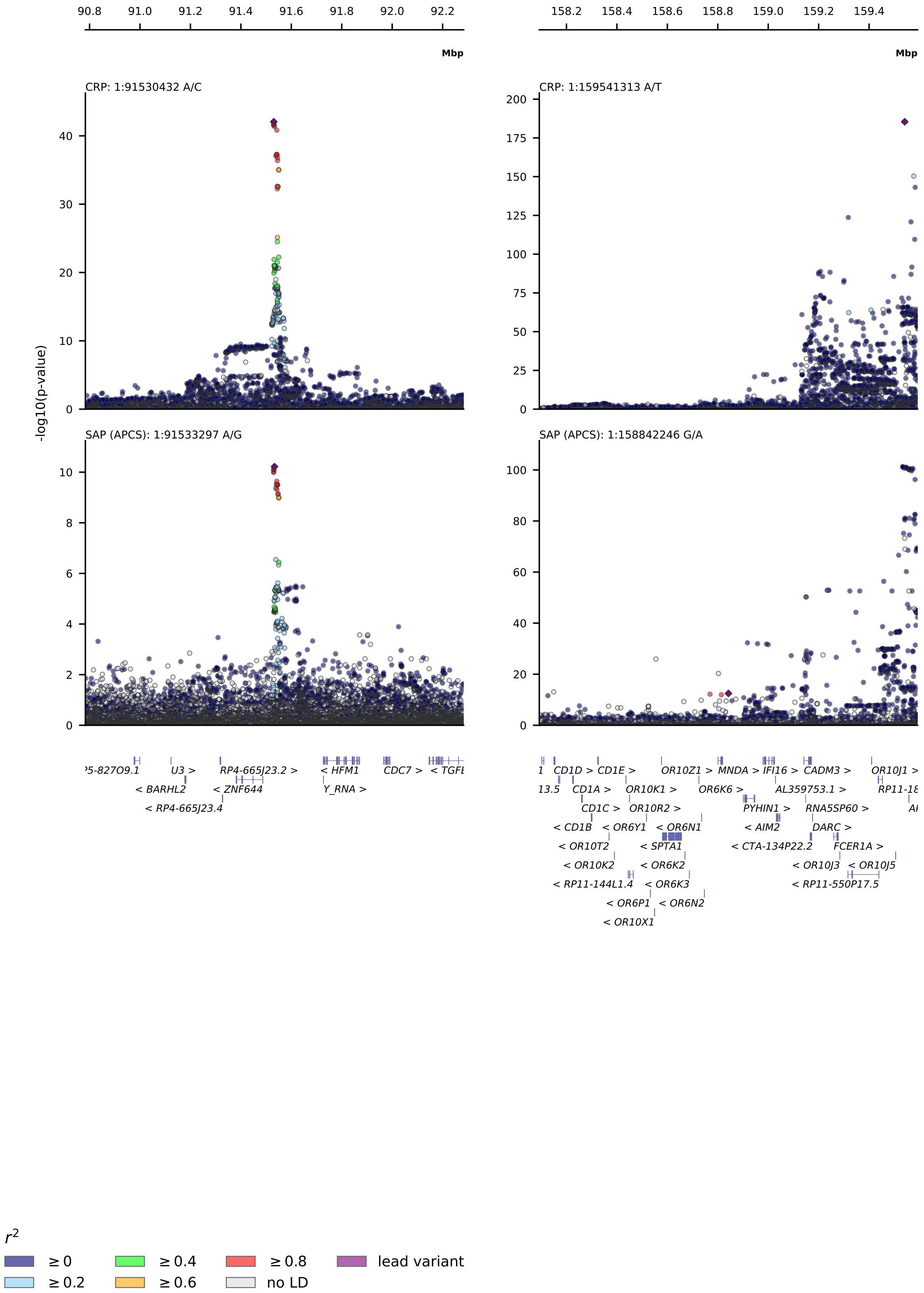
CRP: 1:91530432 A/C
SAP (APCS): 1:91533297 A/G
CRP: 1:159541313 A/T
SAP (APCS): 1:158842246 G/A
-log10(p-value)
Mbp
r2
≥ 0
≥ 0.2
≥ 0.4
≥ 0.6
≥ 0.8
no LD
lead variant

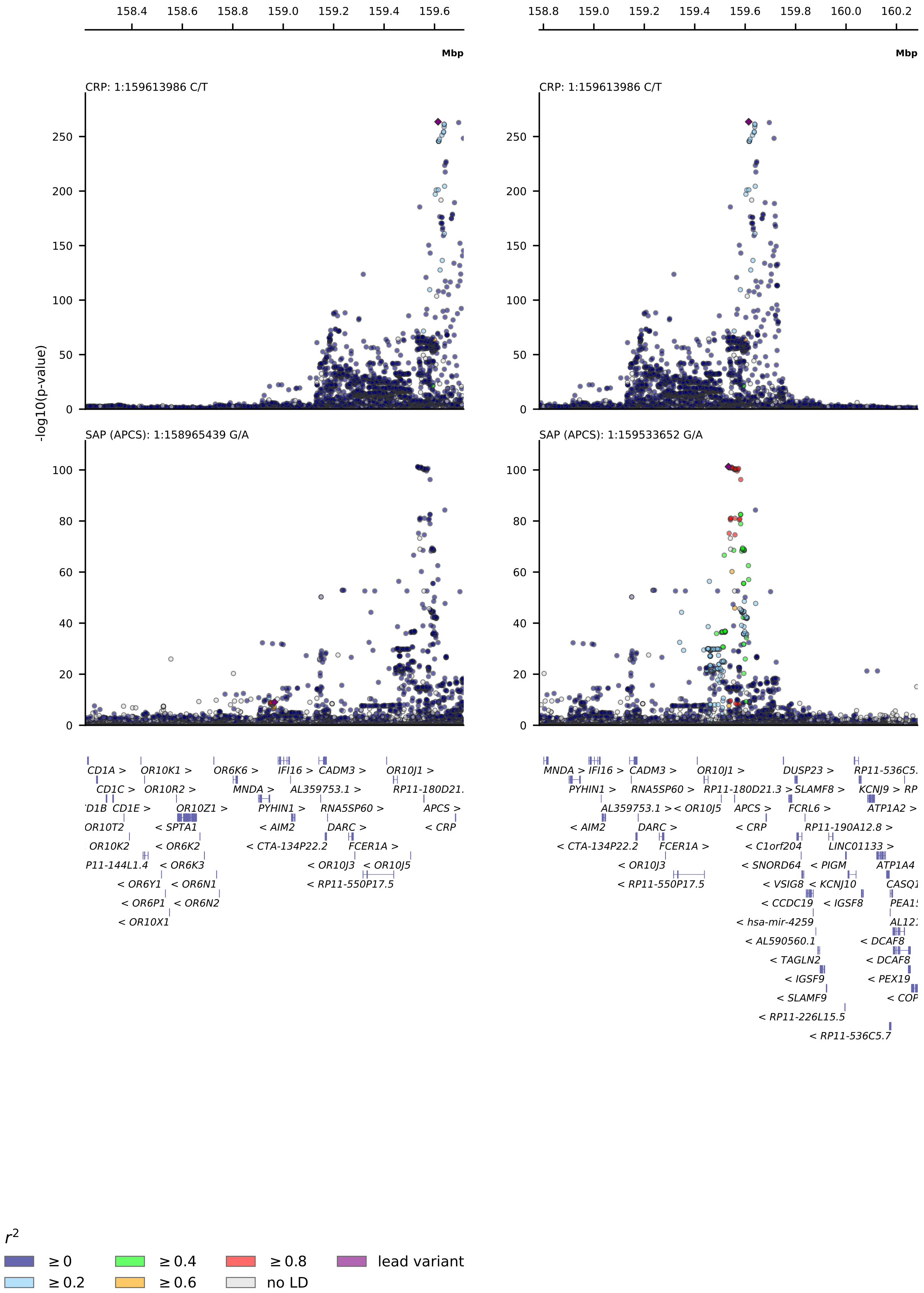
Mbp
CRP: 1:159613986 C/T
SAP (APCS): 1:158965439 G/A
SAP (APCS): 1:159533652 G/A
-log10(p-value)
$r^2$
≥ 0
≥ 0.2
≥ 0.4
≥ 0.6
≥ 0.8
no LD
lead variant

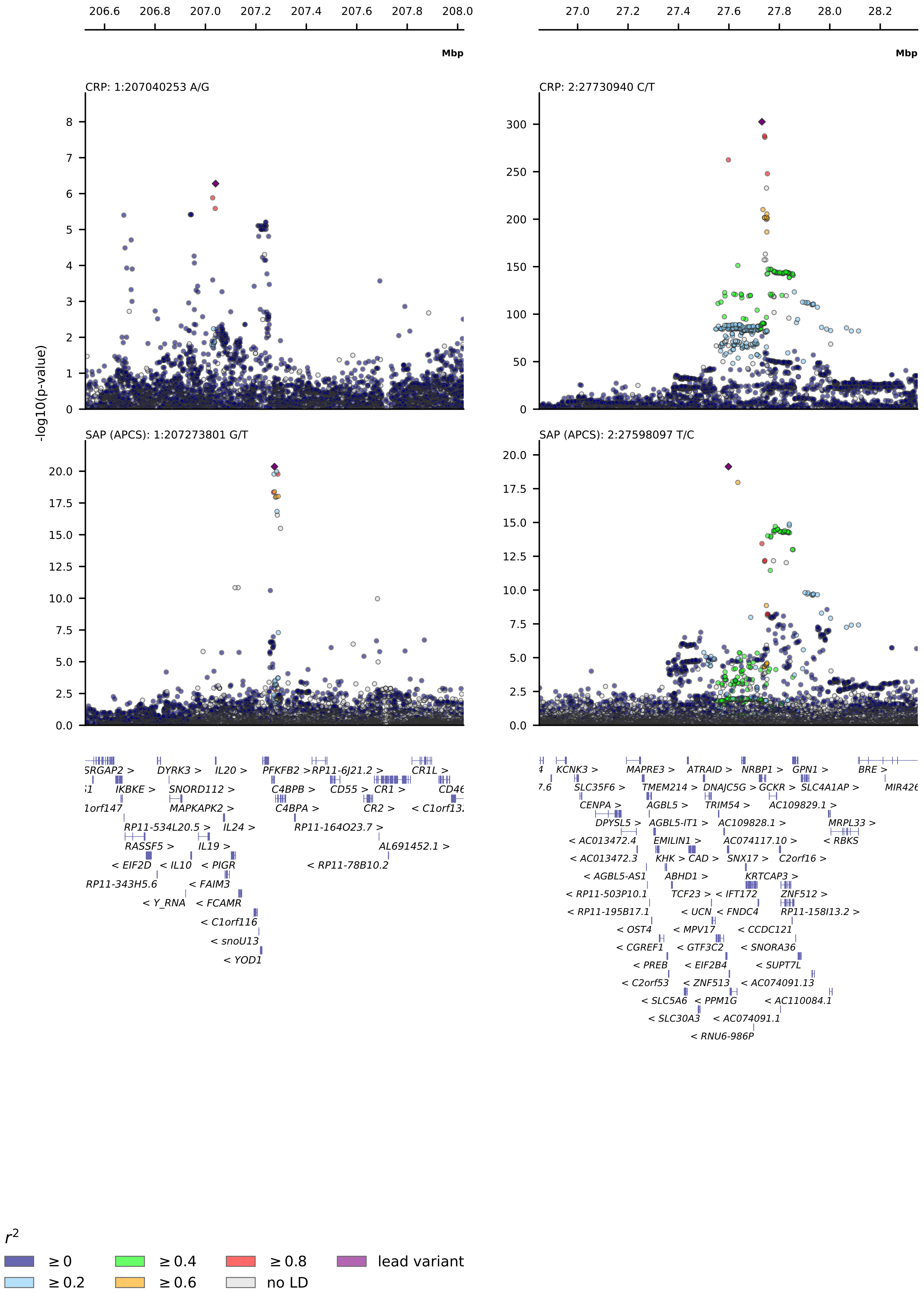
206.6 206.8 207.0 207.2 207.4 207.6 207.8 208.0
Mbp
27.0 27.2 27.4 27.6 27.8 28.0 28.2
Mbp
CRP: 1:207040253 A/G
CRP: 2:27730940 C/T
SAP (APCS): 1:207273801 G/T
SAP (APCS): 2:27598097 T/C
-log10(p-value)
$r^2$
≥ 0
≥ 0.2
≥ 0.4
≥ 0.6
≥ 0.8
no LD
lead variant

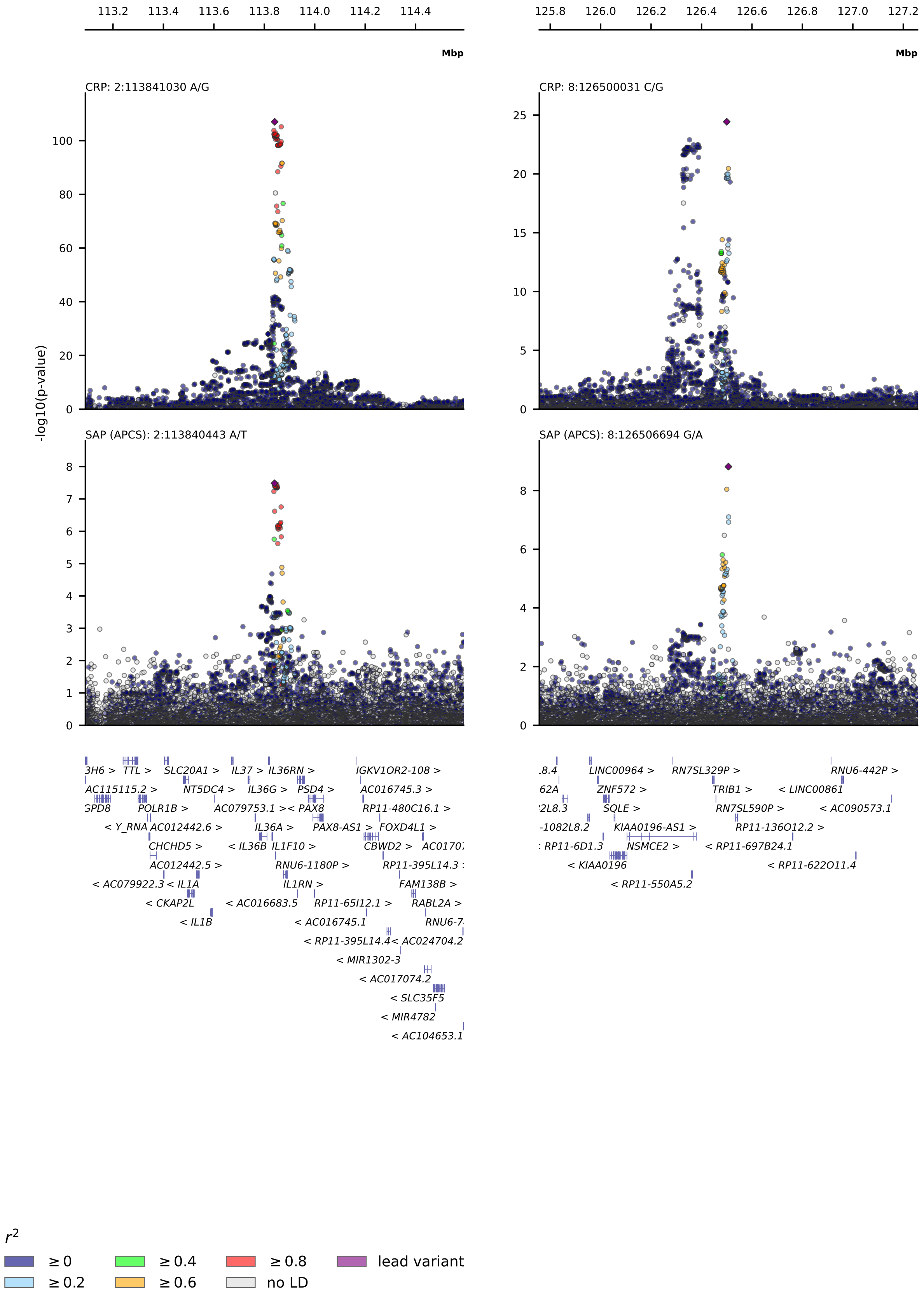
113.2 113.4 113.6 113.8 114.0 114.2 114.4
Mbp
125.8 126.0 126.2 126.4 126.6 126.8 127.0 127.2
Mbp
CRP: 2:113841030 A/G
CRP: 8:126500031 C/G
SAP (APCS): 2:113840443 A/T
SAP (APCS): 8:126506694 G/A
-log10(p-value)
$r^2$
≥ 0
≥ 0.2
≥ 0.4
≥ 0.6
≥ 0.8
no LD
lead variant

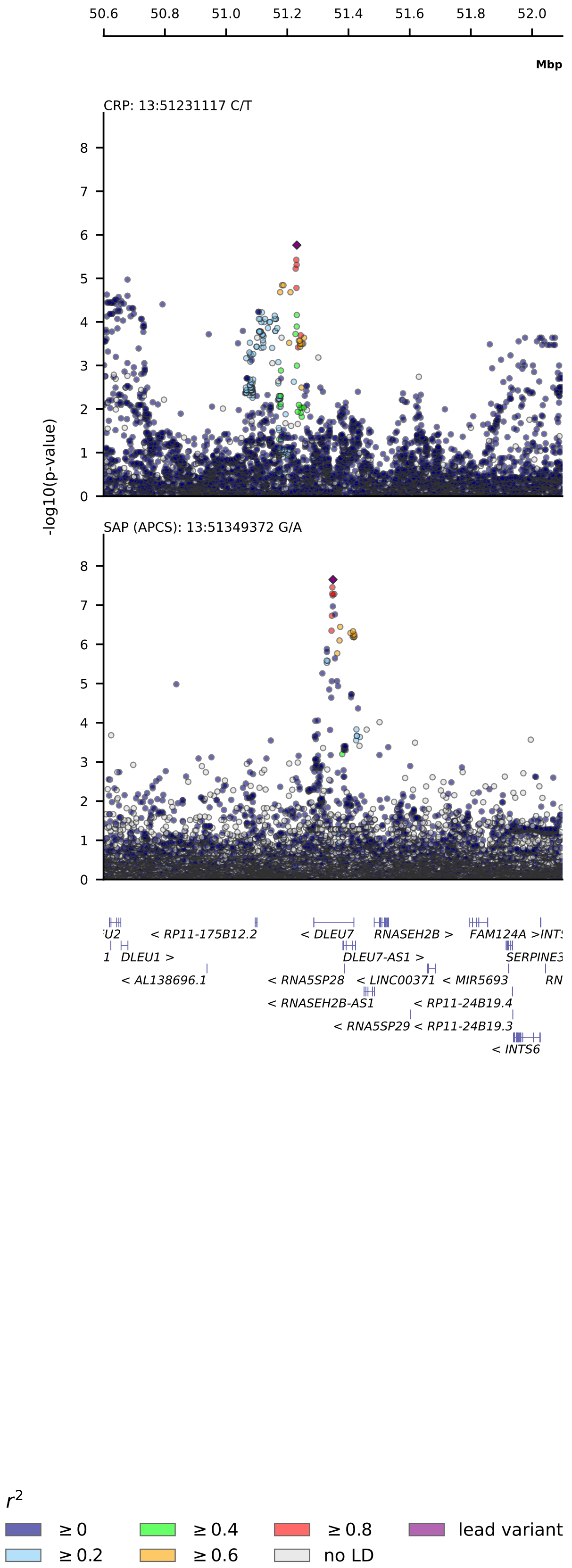
50.6
50.8
51.0
51.2
51.4
51.6
51.8
52.0
Mbp
CRP: 13:51231117 C/T
-log10(p-value)
SAP (APCS): 13:51349372 G/A
'U2
< RP11-175B12.2
< DLEU7
RNASEH2B >
FAM124A
>INTS
1
DLEU1 >
DLEU7-AS1 >
SERPINE3
< AL138696.1
< RNA5SP28
< LINC00371
< MIR5693
RN
< RNASEH2B-AS1
< RP11-24B19.4
< RNA5SP29
< RP11-24B19.3
< INTS6
$r^2$
≥ 0
≥ 0.4
≥ 0.8
lead variant
≥ 0.2
≥ 0.6
no LD

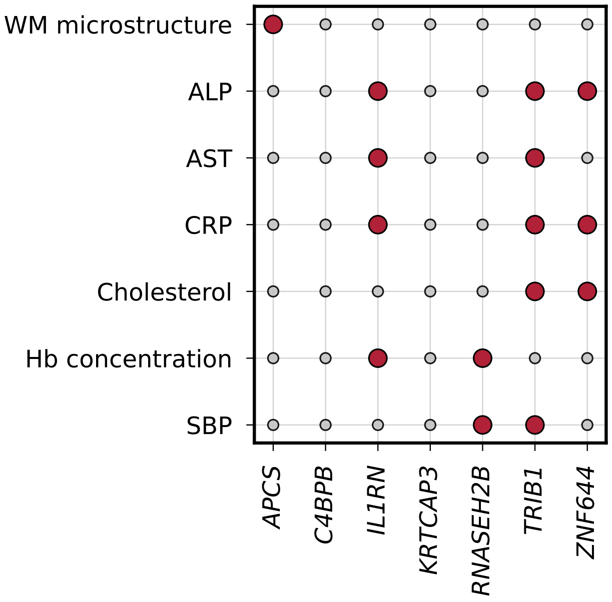

**Figure S3:** GWAS findings for the 'author assigned genes' reported by GWAS Catalog, showing all traits assigned to *APCS* and traits reported for 2 or more of the trans signals. Abbreviations: WM, white matter; Hb, blood haemoglobin concentration; SBP, systolic blood pressure; plasma concentrations of ALP, alkaline phosphatase; AST, aspartate transaminase; CRP, C-reactive protein. Table S9 shows the complete GWAS Catalog look-up.

#### Tables

**Table S1:** Lead variants for plasma serum amyloid P component (SAP) value based on a meta-analysis GWAS in 44,288 subjects. Lead variants from a *trans*-region, defined as a signal beyond +/-2MB with respect to *APCS*, were mapped to putative causal genes using Open Target's 'v2g' pipeline. The provided genetic information is based on genomic build 37.

| lead variant (rsid) | chromosome | start position | effect allele | other allele | effect size | standard error | $-\log_{10}$(p-value) | Nearest protein coding gene | Ensembl id (Nearest protein coding gene) | v2g mapped gene | Ensembl id (v2g mapped gene) |
|---|---|---|---|---|---|---|---|---|---|---|---|
| rs140308485 | 1 | 158842246 | A | G | -0.1903 | 0.0261 | 12.468 | MNDA | ENSG00000163563 | APCS | ENSG00000132703 |
| rs13374652 | 1 | 158965439 | A | G | -0.0892 | 0.0146 | 9.045 | IFI16 | ENSG00000163565 | APCS | ENSG00000132703 |
| rs1341664 | 1 | 159533652 | A | G | -0.2144 | 0.0100 | 101.296 | APCS | ENSG00000132703 | APCS | ENSG00000132703 |
| rs78228389 | 1 | 159783225 | A | G | 0.1335 | 0.0227 | 8.382 | FCRL6 | ENSG00000181036 | APCS | ENSG00000132703 |
| rs2808467 | 1 | 207273801 | G | T | -0.0938 | 0.0100 | 20.363 | C4BPB | ENSG00000123843 | C4BPB | ENSG00000123843 |
| rs165316 | 1 | 91533297 | A | G | 0.0564 | 0.0086 | 10.219 | ZNF644 | ENSG00000122482 | ZNF644 | ENSG00000122482 |
| rs10188292 | 2 | 113840443 | A | T | -0.0395 | 0.0071 | 7.484 | IL1F10 | ENSG00000136697 | IL1RN | ENSG00000136689 |
| rs4665972 | 2 | 27598097 | C | T | -0.0807 | 0.0088 | 19.132 | SNX17 | ENSG00000115234 | KRTCAP3 | ENSG00000157992 |
| rs112875651 | 8 | 126506694 | A | G | -0.0443 | 0.0073 | 8.815 | TRIB1 | ENSG00000173334 | TRIB1 | ENSG00000173334 |
| rs9591359 | 13 | 51349372 | A | G | -0.0517 | 0.0092 | 7.650 | DLEU1 | ENSG00000176124 | RNASEH2B | ENSG00000136104 |

**Table S2:** SAP lead variant associations with plasma concentration of C-reactive protein (ref: 62)

| lead variant (rsid) | chromosome | start position | effect allele | other allele | effect size | standard error | $-\log_{10}$(p-value) |
|---|---|---|---|---|---|---|---|
| rs140308485 | 1 | 158842246 | A | G | -0.0150 | 0.0081 | 1.1836 |
| rs13374652 | 1 | 158965439 | A | G | 0.0005 | 0.0043 | 0.0424 |
| rs1341664 | 1 | 159533652 | A | G | -0.0153 | 0.0031 | 6.2623 |
| rs78228389 | 1 | 159783225 | A | G | -0.0242 | 0.0058 | 4.5163 |
| rs2808467 | 1 | 207273801 | G | T | -0.0028 | 0.0027 | 0.5109 |
| rs165316 | 1 | 91533297 | A | G | 0.0346 | 0.0025 | 41.3587 |
| rs10188292 | 2 | 113840443 | A | T | -0.0439 | 0.0020 | 102.3605 |
| rs4665972 | 2 | 27598097 | C | T | -0.0741 | 0.0021 | 262.4584 |
| rs112875651 | 8 | 126506694 | A | G | -0.0202 | 0.0021 | 20.4605 |
| rs9591359 | 13 | 51349372 | A | G | -0.0017 | 0.0026 | 0.2834 |

**Table S3:** Open target v2g results mapping *trans*-variant rs2808467 to a putative causal gene

| Gene | Overall V2G | Distance (Canonical TSS) | pQTL | sQTL | eQTL | Enhancer-TSS corr (FANTOM5) | PCHi-C (Javierre, 2016) | DHS-promoter corr (Thurman, 2012) | PCHi-C (Jung, 2019) | VEP (Ensembl) |
|---|---|---|---|---|---|---|---|---|---|---|
| *YOD1* | 0.0598 | 47476 | - | - | - | - | - | - | - | - |
| *CD55* | 0.1793 | 221063 | - | - | 0.6 | - | 0.9 | - | - | - |
| *PIGR* | 0.0465 | 153990 | - | - | - | - | - | - | - | - |
| *IL20* | 0.0332 | 234833 | - | - | - | - | - | - | - | - |
| *DYRK3* | 0.0066 | 464920 | - | - | - | - | - | - | - | - |
| *MAPKAPK2* | 0.0133 | 415551 | - | - | - | - | - | - | - | - |
| *C4BPA* | 0.1992 | 3777 | - | - | 1 | - | - | - | - | - |
| *EIF2D* | 0.0000 | 487991 | - | - | - | - | - | - | - | - |
| *IL24* | 0.0398 | 203013 | - | - | - | - | - | - | - | - |
| *IL19* | 0.0199 | 329692 | - | - | - | - | - | - | - | - |
| *FCMR* | 0.0398 | 177209 | - | - | - | - | - | - | - | - |
| *C1orf116* | 0.0598 | 67700 | - | - | - | - | - | - | - | - |
| *C4BPB* | 0.2676 | 11596 | - | 1 | - | - | - | - | - | - |
| *CD46* | 0.0000 | - | - | 0 | - | - | - | - | - | - |
| *IL10* | 0.0199 | 325915 | - | - | - | - | - | - | - | - |
| *CR1* | 0.0598 | 395691 | - | - | - | - | 0.7 | - | - | - |
| *PFKFB2* | 0.0598 | 66090 | - | - | - | - | - | - | - | - |
| *FCAMR* | 0.0465 | 129831 | - | - | - | - | - | - | - | - |
| *CR2* | 0.0531 | 352568 | - | - | - | - | 0.5 | - | - | - |

*General:*
Entries without information are marked by '-'.

**Table S4:** Open target v2g results mapping *trans*-variant rs165316 to a putative causal gene

| Gene | Overall V2G | Distance (Canonical TSS) | pQTL | sQTL | eQTL | Enhancer-TSS corr (FANTOM5) | PCHi-C (Javierre, 2016) | DHS-promoter corr (Thurman, 2012) | PCHi-C (Jung, 2019) | VEP (Ensembl) |
|---|---|---|---|---|---|---|---|---|---|---|
| *ZNF644* | 0.0598 | 45468 | - | - | - | - | - | - | - | - |
| *BARHL2* | 0.0199 | 350438 | - | - | - | - | - | - | - | - |
| *CDC7* | 0.0066 | 433111 | - | - | - | - | - | - | - | - |
| *HFM1* | 0.0199 | 337116 | - | - | - | - | - | - | - | - |

*General:*
Entries without information are marked by '-'.

#### **Table S5:** Open target v2g results mapping *trans*-variant rs10188292 to a putative causal gene

| Gene | Overall V2G | Distance (Canonical TSS) | pQTL | sQTL | eQTL | Enhancer-TSS corr (FANTOM5) | PCHi-C (Javierre, 2016) | DHS-promoter corr (Thurman, 2012) | PCHi-C (Jung, 2019) | VEP (Ensembl) |
|---|---|---|---|---|---|---|---|---|---|---|
| *IL37* | 0.0465 | 171701 | - | - | - | - | - | - | - | - |
| *CBWD2* | 0.0199 | 354825 | - | - | - | - | - | - | - | - |
| *PAX8* | 0.0996 | 196055 | - | - | - | - | 0.9 | - | - | - |
| *IL1F10* | 0.0930 | 14896 | - | - | 0.2 | - | - | - | - | - |
| *IGKV1OR2-108* | 0.0266 | 323530 | - | - | - | - | - | - | - | - |
| *IL36G* | 0.0531 | 109663 | - | - | - | - | - | - | - | - |
| *FOXD4L1* | 0.0133 | 415799 | - | - | - | - | - | - | - | - |
| *CHCHD5* | 0.0000 | 498626 | - | - | - | - | - | - | - | - |
| *IL1B* | 0.0332 | 246050 | - | - | - | - | - | - | - | - |
| *IL36A* | 0.0531 | 77407 | - | - | - | - | - | - | - | - |
| *CKAP2L* | 0.0266 | 318202 | - | - | - | - | - | - | - | - |
| *IL1A* | 0.0266 | 298373 | - | - | - | - | - | - | - | - |
| *IL36RN* | 0.0664 | 24228 | - | - | - | - | - | - | 0 | - |
| *SLC20A1* | 0.0465 | 436927 | - | - | 0.3 | - | - | - | - | - |
| *PSD4* | 0.2058 | 74459 | - | - | 0.7 | - | 0.8 | - | 0 | - |
| *IL36B* | 0.0598 | 29999 | - | - | - | - | - | - | 0 | - |
| *IL1RN* | 0.4058 | 16449 | 0.9 | 0.4 | 0.6 | - | 0.9 | - | - | - |
| *NT5DC4* | 0.0199 | 361846 | - | - | - | - | - | - | - | - |

*General:*
Entries without information are marked by '-'.

**Table S6:** Open target v2g results mapping *trans*-variant rs4665972 to a putative causal gene

| Gene | Overall V2G | Distance (Canonical TSS) | pQTL | sQTL | eQTL | Enhancer-TSS corr (FANTOM5) | PCHi-C (Javierre, 2016) | DHS-promoter corr (Thurman, 2012) | PCHi-C (Jung, 2019) | VEP (Ensembl) |
|---|---|---|---|---|---|---|---|---|---|---|
| *DNAJC5G* | 0.0531 | 99797 | - | - | - | - | - | - | - | - |
| *PREB* | 0.0332 | 240564 | - | - | - | - | - | - | - | - |
| *KHK* | 0.0398 | 288483 | - | - | - | - | 0.2 | - | - | - |
| *TRIM54* | 0.1738 | 92801 | - | 0.6 | - | - | - | - | - | - |
| *MAPRE3* | 0.0133 | 404593 | - | - | - | - | - | - | - | - |
| *GTF3C2* | 0.1328 | 18231 | - | - | - | - | 1 | - | - | - |
| *KRTCAP3* | 0.2799 | 67136 | - | 0.5 | 0.9 | - | - | - | - | - |
| *SNX17* | 0.1994 | 4734 | - | - | 0.8 | - | 0.1 | - | - | intron_variant |
| *ZNF512* | 0.0398 | 207739 | - | - | - | - | - | - | - | - |
| *GPN1* | 0.1531 | 253017 | - | 0.2 | 0.6 | - | 0 | - | - | - |
| *EIF2B4* | 0.1262 | 4892 | - | - | 0.4 | - | 0.1 | - | - | - |
| *PPM1G* | 0.1527 | 34361 | - | - | 0.7 | - | - | - | - | - |
| *IFT172* | 0.2475 | 114575 | - | 0.9 | 0.1 | - | - | - | - | - |
| *ZNF513* | 0.1062 | 5560 | - | - | 0.3 | - | - | - | - | - |
| *NRBP1* | 0.1793 | 52560 | - | - | 0.9 | - | - | - | - | - |
| *TMEM214* | 0.0664 | 342320 | - | - | - | - | 0.7 | - | - | - |
| *SLC4A1AP* | 0.0465 | 288196 | - | - | - | - | 0.3 | - | - | - |
| *CGREF1* | 0.0332 | 256102 | - | - | - | - | - | - | - | - |
| *FNDC4* | 0.2736 | 119970 | - | 0.7 | 0.6 | - | - | - | - | - |
| *ABHD1* | 0.0332 | 251441 | - | - | - | - | - | - | - | - |
| *C2orf16* | 0.0465 | 162156 | - | - | - | - | - | - | - | - |
| *SLC30A3* | 0.0531 | 99413 | - | - | - | - | - | - | - | - |
| *MRPL33* | 0.0133 | 396487 | - | - | - | - | - | - | - | - |
| *AGBL5* | 0.0199 | 332866 | - | - | - | - | - | - | - | - |
| *SLC5A6* | 0.1728 | 162272 | - | 0.1 | 0.8 | - | - | - | - | - |
| *TCF23* | 0.0332 | 226226 | - | - | - | - | - | - | - | - |
| *OST4* | 0.0266 | 303576 | - | - | - | - | 0 | - | - | - |
| *ATRAID* | 0.1527 | 163189 | - | - | 0.8 | - | - | - | - | - |
| *UCN* | 0.0598 | 66785 | - | - | - | - | - | - | - | - |
| *CCDC121* | 0.0332 | 253782 | - | - | - | - | 0 | - | - | - |
| *PRR30* | 0.0332 | 235820 | - | - | - | - | - | - | - | - |
| *SUPT7L* | 0.0465 | 288610 | - | - | - | - | 0.3 | - | - | - |
| *EMILIN1* | 0.0398 | 296615 | - | - | - | - | 0.2 | - | - | - |
| *CAD* | 0.0465 | 157861 | - | - | - | - | - | - | - | - |
| *MPV17* | 0.0598 | 49550 | - | - | - | - | - | - | - | - |
| *GCKR* | 0.2133 | 121609 | - | 0.4 | 0.6 | - | - | - | - | - |

*General:*
Entries without information are marked by '-'.

**Table S7:** Open target v2g results mapping *trans*-variant rs112875651 to a putative causal gene

| Gene | Overall V2G | Distance (Canonical TSS) | pQTL | sQTL | eQTL | Enhancer-TSS corr (FANTOM5) | PCHi-C (Javierre, 2016) | DHS-promoter corr (Thurman, 2012) | PCHi-C (Jung, 2019) | VEP (Ensembl) |
|---|---|---|---|---|---|---|---|---|---|---|
| *SQLE* | 0.0000 | 495955 | - | - | - | - | - | - | - | - |
| *TRIB1* | 0.0598 | 64094 | - | - | - | - | - | - | - | - |
| *WASHC5* | 0.0398 | 402633 | - | - | - | - | 0.4 | - | - | - |
| *NSMCE2* | 0.0398 | 402773 | - | - | - | - | 0.4 | - | - | - |

*General:*
Entries without information are marked by '-'.

**Table S8:** Open target v2g results mapping *trans*-variant rs9591359 to a putative causal gene

| Gene | Overall V2G | Distance (Canonical TSS) | pQTL | sQTL | eQTL | Enhancer-TSS corr (FANTOM5) | PCHi-C (Javierre, 2016) | DHS-promoter corr (Thurman, 2012) | PCHi-C (Jung, 2019) | VEP (Ensembl) |
|---|---|---|---|---|---|---|---|---|---|---|
| *DLEU7* | 0.0799 | 68703 | - | - | - | - | - | - | - | intron_variant |
| *C13orf42* | 0.0066 | 425016 | - | - | - | - | - | - | - | - |
| *RNASEH2B* | 0.0863 | 134511 | - | - | 0.3 | - | - | - | 0 | - |
| *FAM124A* | 0.0066 | 447098 | - | - | - | - | - | - | - | - |

*General:*
Entries without information are marked by '-'.

**Table S9:** GWAS Catalog look-ups for putative causal genes for plasma SAP value.

| Trait | APCS | C4BPB | IL1RN | KRTCAP3 | RNASEH2B | TRIB1 | ZNF644 | Counts |
|---|---|---|---|---|---|---|---|---|
| AAA | 0 | 0 | 0 | 0 | 0 | 1 | 0 | 1 |
| ALP | 0 | 0 | 1 | 0 | 0 | 1 | 1 | 3 |
| ALT | 0 | 0 | 0 | 0 | 0 | 1 | 0 | 1 |
| AST | 0 | 0 | 1 | 0 | 0 | 1 | 0 | 2 |
| Adiponectin | 0 | 0 | 0 | 0 | 0 | 1 | 0 | 1 |
| Albuminuria | 0 | 0 | 0 | 0 | 0 | 1 | 0 | 1 |
| Alcohol consumption | 0 | 0 | 0 | 0 | 0 | 1 | 0 | 1 |
| Apo-A1 | 0 | 0 | 0 | 0 | 0 | 1 | 0 | 1 |
| Apo-B | 0 | 0 | 0 | 0 | 0 | 1 | 0 | 1 |
| BMI | 0 | 0 | 0 | 0 | 0 | 1 | 0 | 1 |
| BPH | 0 | 0 | 0 | 0 | 1 | 0 | 0 | 1 |
| Body height | 0 | 0 | 0 | 0 | 0 | 1 | 0 | 1 |
| CRP | 0 | 0 | 1 | 0 | 0 | 1 | 1 | 3 |
| Cholesterol | 0 | 0 | 0 | 0 | 0 | 1 | 1 | 2 |
| Ferritin | 0 | 0 | 0 | 0 | 0 | 1 | 0 | 1 |
| Fibrinogen | 0 | 0 | 1 | 0 | 0 | 0 | 0 | 1 |
| GGT | 0 | 0 | 0 | 0 | 0 | 1 | 0 | 1 |
| Glycine | 0 | 0 | 0 | 0 | 0 | 1 | 0 | 1 |
| Granulocytes | 0 | 0 | 1 | 0 | 0 | 0 | 0 | 1 |
| HDL-C | 0 | 0 | 0 | 0 | 0 | 1 | 0 | 1 |
| Hb concentration | 0 | 0 | 1 | 0 | 1 | 0 | 0 | 2 |
| HbA1c | 0 | 0 | 0 | 0 | 0 | 1 | 0 | 1 |
| IBD | 0 | 0 | 0 | 0 | 0 | 1 | 0 | 1 |
| IL6 concentration | 0 | 0 | 1 | 0 | 0 | 0 | 0 | 1 |
| Intelligence | 0 | 0 | 0 | 0 | 0 | 0 | 1 | 1 |
| LDL-C | 0 | 0 | 0 | 0 | 0 | 1 | 0 | 1 |
| Lymphocytes | 0 | 0 | 1 | 0 | 0 | 0 | 0 | 1 |
| Metabolic syndrome | 0 | 0 | 0 | 0 | 0 | 1 | 0 | 1 |
| Monocytes | 0 | 0 | 1 | 0 | 0 | 0 | 0 | 1 |
| Neutrophils | 0 | 0 | 1 | 0 | 0 | 0 | 0 | 1 |
| RDW | 0 | 0 | 1 | 0 | 0 | 0 | 0 | 1 |
| SBP | 0 | 0 | 0 | 0 | 1 | 1 | 0 | 2 |
| T2DM | 0 | 0 | 0 | 0 | 0 | 1 | 0 | 1 |
| UTI | 0 | 0 | 0 | 0 | 1 | 0 | 0 | 1 |
| Urate | 0 | 0 | 0 | 0 | 0 | 0 | 1 | 1 |
| Urine alb/creat | 0 | 0 | 0 | 0 | 0 | 1 | 0 | 1 |
| WM microstructure | 1 | 0 | 0 | 0 | 0 | 0 | 0 | 1 |
| eGFR | 0 | 0 | 0 | 0 | 0 | 1 | 0 | 1 |

**Table S10:** The *cis*-Mendelian randomization results for the effects of one standard deviation higher plasma SAP value or plasma CRP concentration on dementia outcomes.

| GWAS pQTL | Exposure (unit) | Outcome trait | Point estimate (95%CI) | No variants | model | Heterogeneity p-value | Heterogeneity chi-square statistic | p-value (scientific notation) | Unit | Multiplicity threshold |
|---|---|---|---|---|---|---|---|---|---|---|
| Interval (n: 3,301) | SAP (SD) | Alzheimer’s disease | 1.08 (1.03; 1.14) | 11 | IVW | 0.0443 | 18.698 | $3.5\times10^{-3}$ | OR | 0.0028 |
| AGES (n: 5,368) | SAP (SD) | Alzheimer’s disease | 1.19 (1.07; 1.33) | 28 | MR Egger | 0.0143 | 44.212 | $1.4\times10^{-3}$ | OR | 0.0028 |
| DECODE (n: 35,559) | SAP (SD) | Alzheimer’s disease | 1.05 (1.00; 1.10) | 58 | IVW | 0.0009 | 96.355 | $6.1\times10^{-2}$ | OR | 0.0028 |
| Combined SL (n: 44,288) | SAP (SD) | Alzheimer’s disease | 1.07 (1.02; 1.11) | 53 | IVW | 0.0002 | 96.500 | $1.8\times10^{-3}$ | OR | 0.0028 |
| CRP (n: 575,531) | CRP (SD) | Alzheimer’s disease | 1.04 (0.94; 1.16) | 132 | MR Egger | 0.0013 | 183.930 | $4.7\times10^{-1}$ | OR | 0.0028 |
| Interval (n: 3,301) | SAP (SD) | Lewy body dementia | 1.57 (1.16; 2.12) | 12 | MR Egger | 0.4782 | 9.579 | $3.1\times10^{-3}$ | OR | 0.0028 |
| AGES (n: 5,368) | SAP (SD) | Lewy body dementia | 1.27 (0.94; 1.72) | 25 | MR Egger | 0.1004 | 31.986 | $1.2\times10^{-1}$ | OR | 0.0028 |
| DECODE (n: 35,559) | SAP (SD) | Lewy body dementia | 0.91 (0.73; 1.14) | 52 | MR Egger | 0.3687 | 52.737 | $4.2\times10^{-1}$ | OR | 0.0028 |
| Combined SL (n: 44,288) | SAP (SD) | Lewy body dementia | 1.37 (1.19; 1.59) | 45 | IVW | 0.3367 | 47.374 | $1.5\times10^{-5}$ | OR | 0.0028 |
| CRP (n: 575,531) | CRP (SD) | Lewy body dementia | 0.82 (0.63; 1.06) | 127 | MR Egger | 0.0000 | 226.145 | $1.2\times10^{-1}$ | OR | 0.0028 |

*General:*

Studies: Interval (ref: 24), DECODE (ref: 25), AGES (ref: 26), Combined SL, total subjects in the three GWAS with SomaLogic values for SAP.

**Table S11:** The *cis*-Mendelian randomization results for the effects of one standard deviation higher plasma SAP value on secondary outcomes

| GWAS pQTL | Exposure (unit) | Outcome trait | Point estimate (95%CI) | No variants | model | Heterogeneity p-value | Heterogeneity chi-square statistic | p-value (scientific notation) | Unit | Multiplicity threshold |
|---|---|---|---|---|---|---|---|---|---|---|
| Interval (n: 3,301) | SAP (SD) | Osteoarthritis | 1.07 (1.03; 1.12) | 13 | MR Egger | 0.0049 | 26.819 | $8.6\times10^{-4}$ | OR | 0.0028 |
| AGES (n: 5,368) | SAP (SD) | Osteoarthritis | 1.01 (1.00; 1.03) | 27 | IVW | 0.1545 | 33.267 | $1.3\times10^{-1}$ | OR | 0.0028 |
| DECODE (n: 35,559) | SAP (SD) | Osteoarthritis | 1.09 (1.06; 1.12) | 58 | MR Egger | 0.0000 | 115.978 | $8.0\times10^{-9}$ | OR | 0.0028 |
| Combined SL (n: 44,288) | SAP (SD) | Osteoarthritis | 1.01 (1.00; 1.03) | 49 | IVW | 0.0127 | 72.505 | $1.2\times10^{-1}$ | OR | 0.0028 |
| Interval (n: 3,301) | SAP (SD) | Osteoarthritis (hip) | 1.13 (1.04; 1.24) | 13 | MR Egger | 0.9109 | 5.389 | $5.4\times10^{-3}$ | OR | 0.0028 |
| AGES (n: 5,368) | SAP (SD) | Osteoarthritis (hip) | 1.11 (1.01; 1.23) | 28 | MR Egger | 0.1388 | 33.851 | $2.6\times10^{-2}$ | OR | 0.0028 |
| DECODE (n: 35,559) | SAP (SD) | Osteoarthritis (hip) | 1.01 (0.97; 1.05) | 59 | IVW | 0.0014 | 95.624 | $6.6\times10^{-1}$ | OR | 0.0028 |
| Combined SL (n: 44,288) | SAP (SD) | Osteoarthritis (hip) | 1.09 (1.02; 1.17) | 55 | MR Egger | 0.1155 | 65.550 | $1.2\times10^{-2}$ | OR | 0.0028 |
| Interval (n: 3,301) | SAP (SD) | Osteoarthritis (hip and knee) | 1.12 (1.04; 1.21) | 12 | MR Egger | 0.0552 | 17.984 | $3.6\times10^{-3}$ | OR | 0.0028 |
| AGES (n: 5,368) | SAP (SD) | Osteoarthritis (hip and knee) | 1.02 (1.00; 1.04) | 27 | IVW | 0.0000 | 73.906 | $6.5\times10^{-2}$ | OR | 0.0028 |
| DECODE (n: 35,559) | SAP (SD) | Osteoarthritis (hip and knee) | 1.10 (1.05; 1.15) | 53 | MR Egger | 0.0061 | 79.838 | $8.5\times10^{-6}$ | OR | 0.0028 |
| Combined SL (n: 44,288) | SAP (SD) | Osteoarthritis (hip and knee) | 1.00 (0.97; 1.02) | 51 | IVW | 0.0000 | 111.507 | $9.2\times10^{-1}$ | OR | 0.0028 |
| Interval (n: 3,301) | SAP (SD) | Osteoarthritis (knee) | 1.14 (1.04; 1.24) | 12 | MR Egger | 0.0692 | 17.242 | $6.3\times10^{-3}$ | OR | 0.0028 |
| AGES (n: 5,368) | SAP (SD) | Osteoarthritis (knee) | 1.15 (1.08; 1.24) | 26 | MR Egger | 0.0005 | 53.489 | $6.1\times10^{-5}$ | OR | 0.0028 |
| DECODE (n: 35,559) | SAP (SD) | Osteoarthritis (knee) | 1.09 (1.04; 1.15) | 55 | MR Egger | 0.0077 | 81.160 | $6.8\times10^{-4}$ | OR | 0.0028 |
| Combined SL (n: 44,288) | SAP (SD) | Osteoarthritis (knee) | 0.97 (0.94; 1.00) | 49 | IVW | 0.0000 | 103.149 | $4.8\times10^{-2}$ | OR | 0.0028 |
| Interval (n: 3,301) | SAP (SD) | SLE | 0.94 (0.78; 1.13) | 10 | IVW | 0.0818 | 15.350 | $4.8\times10^{-1}$ | OR | 0.0028 |
| AGES (n: 5,368) | SAP (SD) | SLE | 0.91 (0.83; 0.99) | 26 | IVW | 0.0257 | 40.531 | $2.7\times10^{-2}$ | OR | 0.0028 |
| DECODE (n: 35,559) | SAP (SD) | SLE | 0.93 (0.83; 1.04) | 49 | IVW | 0.0051 | 76.845 | $2.0\times10^{-1}$ | OR | 0.0028 |
| Combined SL (n: 44,288) | SAP (SD) | SLE | 0.93 (0.85; 1.01) | 45 | IVW | 0.0015 | 77.066 | $9.5\times10^{-2}$ | OR | 0.0028 |
| Interval (n: 3,301) | SAP (SD) | IPF | 1.00 (1.00; 1.00) | 13 | IVW | 0.0296 | 22.785 | $2.1\times10^{-3}$ | OR | 0.0028 |
| AGES (n: 5,368) | SAP (SD) | IPF | 1.00 (1.00; 1.00) | 28 | MR Egger | 0.5940 | 23.684 | $2.1\times10^{-1}$ | OR | 0.0028 |
| DECODE (n: 35,559) | SAP (SD) | IPF | 1.00 (1.00; 1.00) | 58 | MR Egger | 0.0959 | 70.213 | $1.9\times10^{-1}$ | OR | 0.0028 |
| Combined SL (n: 44,288) | SAP (SD) | IPF | 1.00 (1.00; 1.00) | 52 | IVW | 0.0035 | 82.371 | $7.1\times10^{-1}$ | OR | 0.0028 |
| Interval (n: 3,301) | SAP (SD) | SBP | -0.03 (-0.16; 0.11) | 10 | IVW | 0.3014 | 10.637 | $7.0\times10^{-1}$ | mmHg | 0.0028 |
| AGES (n: 5,368) | SAP (SD) | SBP | -0.08 (-0.33; 0.18) | 27 | MR Egger | 0.1258 | 33.212 | $5.5\times10^{-1}$ | mmHg | 0.0028 |
| DECODE (n: 35,559) | SAP (SD) | SBP | -0.21 (-0.47; 0.06) | 41 | MR Egger | 0.1069 | 50.255 | $1.3\times10^{-1}$ | mmHg | 0.0028 |
| Combined SL (n: 44,288) | SAP (SD) | SBP | -0.16 (-0.26; -0.07) | 46 | IVW | 0.0005 | 82.593 | $1.1\times10^{-3}$ | mmHg | 0.0028 |
| Interval (n: 3,301) | SAP (SD) | DBP | -0.55 (-1.13; 0.04) | 10 | MR Egger | 0.0763 | 14.214 | $6.7\times10^{-2}$ | mmHg | 0.0028 |
| AGES (n: 5,368) | SAP (SD) | DBP | -0.04 (-0.10; 0.01) | 28 | IVW | 0.0044 | 50.121 | $1.2\times10^{-1}$ | mmHg | 0.0028 |
| DECODE (n: 35,559) | SAP (SD) | DBP | 0.03 (-0.04; 0.11) | 46 | IVW | 0.0001 | 88.512 | $3.9\times10^{-1}$ | mmHg | 0.0028 |
| Combined SL (n: 44,288) | SAP (SD) | DBP | -0.09 (-0.15; -0.03) | 44 | IVW | 0.0002 | 84.341 | $2.2\times10^{-3}$ | mmHg | 0.0028 |
| Interval (n: 3,301) | SAP (SD) | CHD | 1.03 (1.01; 1.05) | 13 | IVW | 0.7922 | 7.909 | $3.5\times10^{-4}$ | OR | 0.0028 |
| AGES (n: 5,368) | SAP (SD) | CHD | 1.01 (0.97; 1.05) | 26 | MR Egger | 0.0000 | 62.829 | $7.6\times10^{-1}$ | OR | 0.0028 |
| DECODE (n: 35,559) | SAP (SD) | CHD | 0.99 (0.97; 1.01) | 54 | IVW | 0.0011 | 90.282 | $3.1\times10^{-1}$ | OR | 0.0028 |
| Combined SL (n: 44,288) | SAP (SD) | CHD | 1.03 (1.01; 1.05) | 50 | IVW | 0.0044 | 78.772 | $7.3\times10^{-4}$ | OR | 0.0028 |
| Interval (n: 3,301) | SAP (SD) | T2DM | 1.02 (1.00; 1.05) | 10 | IVW | 0.0131 | 20.896 | $8.8\times10^{-2}$ | OR | 0.0028 |
| AGES (n: 5,368) | SAP (SD) | T2DM | 1.02 (1.00; 1.04) | 26 | IVW | 0.0126 | 43.401 | $3.6\times10^{-2}$ | OR | 0.0028 |
| DECODE (n: 35,559) | SAP (SD) | T2DM | 0.96 (0.92; 1.00) | 50 | MR Egger | 0.0208 | 69.981 | $5.1\times10^{-2}$ | OR | 0.0028 |
| Combined SL (n: 44,288) | SAP (SD) | T2DM | 1.01 (0.98; 1.03) | 49 | IVW | 0.0306 | 67.935 | $6.0\times10^{-1}$ | OR | 0.0028 |
| Interval (n: 3,301) | SAP (SD) | ALT | 0.00 (-0.00; 0.01) | 13 | MR Egger | 0.2146 | 14.343 | $9.9\times10^{-2}$ | log(U/L) | 0.0028 |
| AGES (n: 5,368) | SAP (SD) | ALT | 0.00 (-0.00; 0.00) | 29 | IVW | 0.2893 | 31.643 | $6.7\times10^{-1}$ | log(U/L) | 0.0028 |
| DECODE (n: 35,559) | SAP (SD) | ALT | -0.00 (-0.00; 0.00) | 57 | MR Egger | 0.8466 | 44.375 | $7.7\times10^{-1}$ | log(U/L) | 0.0028 |
| Combined SL (n: 44,288) | SAP (SD) | ALT | 0.00 (-0.00; 0.00) | 52 | IVW | 0.1701 | 60.510 | $1.9\times10^{-1}$ | log(U/L) | 0.0028 |
| Interval (n: 3,301) | SAP (SD) | AST | -0.00 (-0.02; 0.01) | 7 | IVW | 0.8304 | 2.825 | $5.5\times10^{-1}$ | SD | 0.0028 |
| AGES (n: 5,368) | SAP (SD) | AST | -0.00 (-0.01; 0.01) | 16 | IVW | 0.3593 | 16.348 | $9.8\times10^{-1}$ | SD | 0.0028 |
| DECODE (n: 35,559) | SAP (SD) | AST | -0.02 (-0.04; 0.00) | 30 | MR Egger | 0.0023 | 53.883 | $7.7\times10^{-2}$ | SD | 0.0028 |
| Combined SL (n: 44,288) | SAP (SD) | AST | -0.00 (-0.01; 0.00) | 30 | IVW | 0.0020 | 55.860 | $1.4\times10^{-1}$ | SD | 0.0028 |

| | | | | | | | | | | |
|---|---|---|---|---|---|---|---|---|---|---|
| Interval (n: 3,301) | SAP (SD) | GGT | 0.00 (-0.00; 0.00) | 13 | IVW | 0.0060 | 27.737 | $6.0\times10^{-2}$ | log(U/L) | 0.0028 |
| AGES (n: 5,368) | SAP (SD) | GGT | -0.00 (-0.00; 0.00) | 28 | IVW | 0.0129 | 45.937 | $8.8\times10^{-1}$ | log(U/L) | 0.0028 |
| DECODE (n: 35,559) | SAP (SD) | GGT | 0.00 (0.00; 0.00) | 58 | IVW | 0.0001 | 107.573 | $1.5\times10^{-2}$ | log(U/L) | 0.0028 |
| Combined SL (n: 44,288) | SAP (SD) | GGT | 0.00 (0.00; 0.01) | 52 | MR Egger | 0.0002 | 93.710 | $5.0\times10^{-2}$ | log(U/L) | 0.0028 |
| Interval (n: 3,301) | SAP (SD) | Total brain volume | 0.01 (-0.02; 0.04) | 11 | IVW | 0.7042 | 7.223 | $6.3\times10^{-1}$ | SD | 0.0028 |
| AGES (n: 5,368) | SAP (SD) | Total brain volume | 0.01 (-0.01; 0.03) | 30 | IVW | 0.5319 | 27.739 | $1.7\times10^{-1}$ | SD | 0.0028 |
| DECODE (n: 35,559) | SAP (SD) | Total brain volume | -0.01 (-0.04; 0.02) | 50 | IVW | 0.8835 | 37.548 | $5.7\times10^{-1}$ | SD | 0.0028 |
| Combined SL (n: 44,288) | SAP (SD) | Total brain volume | 0.06 (0.02; 0.10) | 50 | MR Egger | 0.3700 | 50.634 | $2.0\times10^{-3}$ | SD | 0.0028 |
| Interval (n: 3,301) | SAP (SD) | Cerebral WMH volume | -0.08 (-0.14; -0.03) | 10 | IVW | 0.3732 | 9.725 | $2.2\times10^{-3}$ | white | 0.0028 |
| AGES (n: 5,368) | SAP (SD) | Cerebral WMH volume | -0.12 (-0.20; -0.04) | 29 | MR Egger | 0.6213 | 24.162 | $4.3\times10^{-3}$ | white | 0.0028 |
| DECODE (n: 35,559) | SAP (SD) | Cerebral WMH volume | -0.06 (-0.10; -0.02) | 55 | IVW | 0.0238 | 76.467 | $3.9\times10^{-3}$ | white | 0.0028 |
| Combined SL (n: 44,288) | SAP (SD) | Cerebral WMH volume | -0.03 (-0.07; 0.01) | 52 | IVW | 0.0411 | 69.816 | $1.4\times10^{-1}$ | white | 0.0028 |
| Interval (n: 3,301) | SAP (SD) | Circulating total tau | 0.03 (-0.00; 0.06) | 11 | IVW | 0.0299 | 19.935 | $6.6\times10^{-2}$ | log2 | 0.0028 |
| AGES (n: 5,368) | SAP (SD) | Circulating total tau | -0.02 (-0.04; 0.01) | 27 | IVW | 0.1325 | 34.101 | $1.7\times10^{-1}$ | log2 | 0.0028 |
| DECODE (n: 35,559) | SAP (SD) | Circulating total tau | 0.05 (0.02; 0.08) | 53 | IVW | 0.0141 | 76.877 | $3.6\times10^{-3}$ | log2 | 0.0028 |
| Combined SL (n: 44,288) | SAP (SD) | Circulating total tau | -0.09 (-0.16; -0.02) | 49 | MR Egger | 0.0126 | 71.314 | $1.8\times10^{-2}$ | log2 | 0.0028 |

*General:*

Studies: Interval (ref: 24), DECODE (ref: 25), AGES (ref: 26), Combined SL, total subjects in the three GWAS with SomaLogic values for SAP. Outcome trait abbreviations: SLE, systemic lupus erythematosus; IPF, idiopathic pulmonary fibrosis, SBP, systolic blood pressure; DBP, diastolic blood pressure; CHD, coronary heart disease; T2DM, type 2 diabetes mellitus; ALT, AST, GGT, plasma concentrations of, respectively, alanine transaminase, aspartate transaminase, $\gamma$-glutamyl transferase; cerebral WMH, white matter hyperintensity, volume.

**Table S12:** The *cis*-Mendelian randomization results for the effects of one standard deviation higher plasma CRP concentration on secondary outcomes

| GWAS pQTL | Exposure (unit) | Outcome trait | Point estimate (95%CI) | No variants | model | Heterogeneity p-value | Heterogeneity chi-square statistic | p-value (scientific notation) | Unit | Multiplicity threshold |
|---|---|---|---|---|---|---|---|---|---|---|
| CRP (n: 575,531) | CRP (SD) | Osteoarthritis | 1.04 (1.00; 1.08) | 153 | MR Egger | 0.0000 | 251.92 | $2.9\times10^{-2}$ | OR | 0.0028 |
| CRP (n: 575,531) | CRP (SD) | Osteoarthritis (hip) | 1.15 (1.08; 1.23) | 172 | MR Egger | 0.0000 | 258.36 | $2.3\times10^{-5}$ | OR | 0.0028 |
| CRP (n: 575,531) | CRP (SD) | Osteoarthritis (hip and knee) | 1.02 (0.98; 1.07) | 158 | MR Egger | 0.0000 | 321.26 | $3.4\times10^{-1}$ | OR | 0.0028 |
| CRP (n: 575,531) | CRP (SD) | Osteoarthritis (knee) | 0.99 (0.94; 1.05) | 165 | MR Egger | 0.0000 | 316.49 | $8.0\times10^{-1}$ | OR | 0.0028 |
| CRP (n: 575,531) | CRP (SD) | SLE | 0.77 (0.62; 0.95) | 110 | MR Egger | 0.0000 | 204.75 | $1.4\times10^{-2}$ | OR | 0.0028 |
| CRP (n: 575,531) | CRP (SD) | IPF | 1.00 (1.00; 1.00) | 166 | MR Egger | 0.0000 | 323.75 | $8.4\times10^{-2}$ | OR | 0.0028 |
| CRP (n: 575,531) | CRP (SD) | SBP | -0.05 (-0.20; 0.11) | 102 | IVW | 0.0000 | 180.05 | $5.5\times10^{-1}$ | mmHg | 0.0028 |
| CRP (n: 575,531) | CRP (SD) | DBP | 0.05 (-0.05; 0.15) | 105 | IVW | 0.0000 | 182.27 | $2.9\times10^{-1}$ | mmHg | 0.0028 |
| CRP (n: 575,531) | CRP (SD) | CHD | 1.02 (0.99; 1.05) | 151 | MR Egger | 0.0000 | 241.24 | $2.2\times10^{-1}$ | OR | 0.0028 |
| CRP (n: 575,531) | CRP (SD) | T2DM | 1.03 (0.99; 1.07) | 140 | MR Egger | 0.0000 | 235.82 | $1.5\times10^{-1}$ | OR | 0.0028 |
| CRP (n: 575,531) | CRP (SD) | ALT | 0.01 (0.00; 0.01) | 165 | IVW | 0.0000 | 340.31 | $4.1\times10^{-8}$ | log(U/L) | 0.0028 |
| CRP (n: 575,531) | CRP (SD) | AST | 0.02 (0.01; 0.03) | 50 | IVW | 0.0010 | 85.16 | $3.3\times10^{-4}$ | SD | 0.0028 |
| CRP (n: 575,531) | CRP (SD) | GGT | -0.00 (-0.00; 0.00) | 156 | IVW | 0.0000 | 284.49 | $8.5\times10^{-1}$ | log(U/L) | 0.0028 |
| CRP (n: 575,531) | CRP (SD) | Total brain volume | 0.04 (0.00; 0.08) | 122 | MR Egger | 0.0001 | 187.06 | $4.7\times10^{-2}$ | SD | 0.0028 |
| CRP (n: 575,531) | CRP (SD) | Cerebral WMH volume | 0.01 (-0.07; 0.09) | 137 | MR Egger | 0.0000 | 214.52 | $8.5\times10^{-1}$ | white | 0.0028 |
| CRP (n: 575,531) | CRP (SD) | Circulating total tau | 0.20 (0.17; 0.24) | 143 | IVW | 0.0002 | 209.08 | $1.0\times10^{-100}$ | log2 | 0.0028 |

*General:*
Outcome trait abbreviations: SLE, systemic lupus erythematosus; IPF, idiopathic pulmonary fibrosis, SBP, systolic blood pressure; DBP, diastolic blood pressure; CHD, coronary heart disease; T2DM, type 2 diabetes mellitus; ALT, AST, GGT, plasma concentrations of, respectively, alanine transaminase, aspartate transaminase, $\gamma$-glutamyl transferase; cerebral WMH, white matter hyperintensity, volume.

**Table S13:** The *cis*-multivariable Mendelian randomization results for the effects of one standard deviation higher plasma SAP value or plasma CRP concentration.

| Exposure \| co-variate | Exposure (unit) | Outcome trait | Point estimate (95%CI) | No variants | model | Heterogeneity p-value | Heterogeneity chi-square statistic | p-value (scientific notation) | Unit | Multiplicity threshold |
|---|---|---|---|---|---|---|---|---|---|---|
| SAP \| CRP | SAP (SD) | Osteoarthritis (hip) | 1.11 (1.04; 1.19) | 30 | MR Egger | 0.6400 | 23.83 | $2.98\times10^{-3}$ | OR | 0.0028 |
| CRP \| SAP | CRP log(mg/L) | Osteoarthritis (hip) | 1.02 (0.93; 1.11) | 30 | IVW | 0.2612 | 32.33 | $6.46\times10^{-1}$ | OR | 0.0028 |
| SAP \| CRP | SAP (SD) | Total brain volume | -0.00 (-0.02; 0.01) | 30 | IVW | 0.8214 | 21.09 | $6.38\times10^{-1}$ | SD | 0.0028 |
| CRP \| SAP | CRP log(mg/L) | Total brain volume | 0.04 (-0.01; 0.09) | 30 | IVW | 0.8214 | 21.09 | $1.11\times10^{-1}$ | SD | 0.0028 |
| SAP \| CRP | SAP (SD) | Circulating total tau | 0.06 (0.03; 0.08) | 29 | IVW | 0.6120 | 24.33 | $4.55\times10^{-6}$ | log2 | 0.0028 |
| CRP \| SAP | CRP log(mg/L) | Circulating total tau | 0.20 (0.14; 0.25) | 29 | IVW | 0.6120 | 24.33 | $2.62\times10^{-12}$ | log2 | 0.0028 |
